# The impact on the mental health and well-being of unpaid carers affected by social distancing, self-isolation and shielding during the COVID 19 pandemic in England – a systematic review

**DOI:** 10.1101/2021.08.20.21262375

**Authors:** Tabo Akafekwa, Elizabeth Dalgarno, Arpana Verma

## Abstract

**Objective:** This study explores the impact of the COVID 19 lockdown measures on the mental health and well-being of unpaid carers, who make up the largest number of the carer population in England.

**Study design:** A systematic review research protocol was designed and used to conduct the review along with the Enhancing Transparency in Reporting the synthesis of Qualitative Research - ENTREQ statement [43]. Pre-determined inclusion and exclusion criteria were used. EndNote X9 reference management was used and the search process was represented using the Preferred Reporting Items for Systematic Reviews and Meta-Analyses (PRISMA) flow diagram [76]. Appraisal of the included research was carried out using the Critical Appraisal Skills Programme (CASP) [57]. Line by line coding was done using inductive thematic synthesis and EPPI Reviewer 4 software [60].

**Results:** Four themes emerged; immediate worries or fears, adapting to change, post pandemic fears and use of technology.

**Conclusion:** The measures put in place during the first lockdown period have had detrimental impacts on unpaid carers, putting them at greater risk of burnout. However, use of digital platforms could have a positive impact on well-being. Recommendations for further research are provided.

**What is new?:** *Key findings?:*

- Discontinued or reduced access to activities and services during the first lockdown during the pandemic has had a negative impact on both people who require care and their carers.
- Carers prioritise the mental health and wellbeing needs of the people they care for over their own.
- Further qualitative research from different groups of carers would be useful to gain a deeper understanding of the impact of the COVID 19 pandemic measures on unpaid carers.
- Use of digital technology and digital platforms may be useful tools for carers both during the pandemic and after.

*What this adds to what is known?:*

- There have been very few qualitative studies on the impact of the COVID 19 pandemic on the mental health and wellbeing of unpaid carers, this review has synthesised their findings and will contribute to future research.
- Unpaid carers are known to be at risk of poor mental health and wellbeing outcomes, this review demonstrates that they are even more at risk due to the increased reliance on them during the pandemic.

*What is this implication and what should change?:*

- There is limited qualitative data available from a range of different groups of carers for example, spouse carers, parent carers, carers of people who have specific needs or conditions. Therefore, purposeful sample research to determine the needs of groups of carers during the COVID 19 pandemic could be valuable.
- Unpaid carers who do not have appropriate support are more at risk of poor mental health and wellbeing outcomes. During the pandemic services have had to adapt to the various rules implemented. Digital adaptations to the provision of support to both carers and the people they care for could be beneficial both during and after the pandemic.

## 1. Background

On the 11th of March 2020, the World Health Organisation (WHO) declared a coronavirus - COVID 19 Pandemic [1]. This prompted a national lockdown on the 23^rd^ of March and social distancing was implemented. Many businesses and venues were closed and gatherings of more than two people were stopped [2]. The concept of shielding was also introduced for people who were clinically extremely vulnerable who needed to self-isolate and take extra precaution to protect themselves from the virus [3].

The word carer will be used throughout this article to refer to adults (18 years old and above) who are providing unpaid or informal home-care to a loved-one, friend, neighbour or other known person [4]. The terms ‘unpaid carer’ and ‘informal carer’ are often disputed; however, the term unpaid carer is utilised by the English government and so has been adopted [5] in accordance with the setting of the study. This care can be offered to people with complex needs as well as to those requiring less intensive help with relatively simple tasks, for example domestic aid for frail older people [6, 7].

The government estimates there are over five million unpaid carers in the UK [8]. In 2016 the Office for National Statistics estimated that unpaid adult carers provided care which would have amounted to health and social care cost expenditure of £56.9 billion if paid carers or nursing assistants were employed - an increase of 45.8% from 2005 [9]. Additionally, Buckner and Yeandle [10] calculate an economic value of carers at £132 billion per year in the UK. This figure is expected to continue increasing due to an aging population and it is likely that three in five people will become carers during their lifetime [11]. In addition to this, growing instability in the formal care sector’s labour market created by the pandemic have increased market fragility, placing greater pressure on informal carers and voluntary agencies, whilst increasing unmet care needs [12]

### 1.1 Current UK government laws, policies and initiatives reflecting the significance of the topic

The Care Act (CA) 2014 legally recognises carers and the role they play; it signifies that carers are legally entitled to assessments and support [13]. The Carers Action Plan 2018 – 2020 [14] was also developed using views of carers and agencies who work with carers to recognise and support carers.

In 2018, a strategy was developed which aims to use existing evidence to understand what loneliness is and the impact it has on health and well-being; becoming a carer is identified as a possible trigger for experiencing loneliness [15].

The NHS Long Term Plan also has a section focussing on the health and well-being of carers and acknowledges that carers are twice as likely to experience poor health as non-carers [16].

### 1.2 Definition of mental health and well-being

‘Mental health is an integral part of health; indeed, there is no health without mental health.’ [17, Internet]. Good mental health is a facilitator to coping with life, realising potential and building and maintaining relationships [18] and results in a well-functioning individual [19]. The term mental health is often linked to the term well-being [17 and 18].

The concept of well-being for both carers and people who require care consists of personal dignity, physical health, mental health and emotional well-being, protection from abuse and neglect and having control of their everyday life [20]. It also includes participation in activities such as work, education, training or recreation, social and economic well-being, family and personal relationships, suitable living accommodation and contributing to society [20].

### 1.3 Mental health and well-being of carers before the COVID 19 pandemic

Physical and emotional exhaustion and mental health problems can be experienced by carers [21, 22, 23] and their quality of life can be affected negatively by taking on a caring role [22, 24]. Family burden is frequently reported and also increases the likelihood of a move to nursing home care for the person being cared for [6, 25, 26].

Over the years there has been an increase in the number of people juggling care and employment, of carers leaving employment to become fulltime carers and increased numbers of people reducing work hours to provide care [27]. A correlation between increased financial difficulties because of their caring role and increased feelings of social isolation as well as reduced contact with people is evident [28]. More recently, in comparison to the general public, carers were seven times more likely to report always or often feeling lonely, for carers with financial difficulties the figure increases tenfold [28]. Additionally, more carers who provided a higher number of hours of care reported feelings of social isolation and of less contact with people [28]. It should be noted however that the number of hours spent caring does not always result in an increased negative impact on mental health and well-being. Factors such as other responsibilities the carer has or the reason for caring can also determine the impact of the caring role [11, 21].

### 1.4 What is known about the mental health and well-being of carers during the COVID 19 pandemic?

In response to the COVID19 pandemic Carers UK and Carers Trust [29] released a joint statement in which they emphasised an increased risk of loneliness and isolation for carers. Concern was also raised about lack of access to social care services which would impact the provision of respite or access to paid carers [29]. Following this, research has revealed that burnout is a concern for 55% of carers, the COVID19 outbreak has caused 70% of carers to increase the care they provide and the closure or reduction of local services means that 35% of carers provide more care [11].

In varying degrees support and respite services were reduced during the first lockdown which resulted in increased reliance on carers [30–33]. This is likely to have a detrimental effect on the health and well-being of carers who are relied on to provide increased care during the pandemic [34].

Elsewhere, grey literature has identified that lockdown measures have had a detrimental impact on carers mental health [30, 31, 35, 36]. However, a minority reported improvements in their mental and emotional health [30] which was echoed by one caller to a radio phone in on the topic [35].

There are currently a limited number of in-depth qualitative research studies which explore the impact of the pandemic on the mental health and well-being of carers, in England and worldwide. [37–41]

### 1.5 Aim

This review aimed to explore the impact of social distancing, social-isolation and shielding; as measures implemented during the first lockdown of the COVID 19 pandemic, on the mental health and well-being of carers in England. The review focused on qualitative research which explored the experience and views of carers during the current COVID 19 pandemic using the research question:

How is the mental health and well-being of unpaid carers in England affected by the current measures in place to reduce the spread of the COVID 19 pandemic? This research aimed to:

Synthesise existing qualitative evidence in this area
Develop recommendations to address the impact upon unpaid carer well-being and
Identify gaps for further research.

## 2 Study Design

A research protocol was developed using the National Institute for Health Research (NIHR) [42] PROSPERO - International prospective register of systematic reviews format (Appendix A) and used to guide this review.

To report this study in a methodical manner the ENTREQ statement [43] (Appendix B) has been used which is widely used as a guideline to report the synthesis of qualitative evidence [44, 45]. However, it has been suggested that the statement still requires validation and may need to be used with some caution due to possible limitations of the guideline [43, 44]. Therefore, two papers from the Cochrane Qualitative and Implementation Methods Group guidance series [46, 47]and Noyes et al. [48] chapter on qualitative evidence have also been used.

### 2.1 Search approach

A scoping search identified a gap in evidence about the impact of COVID 19 on the mental health and well-being of carers and confirmed a similar systematic review had not been carried out.

The search strategy and inclusion criteria (Table 1) were built around the research question which was formulated using Population, phenomenon of Interest and Context (PICo) [49, 50] as follows:

**Table 1.**
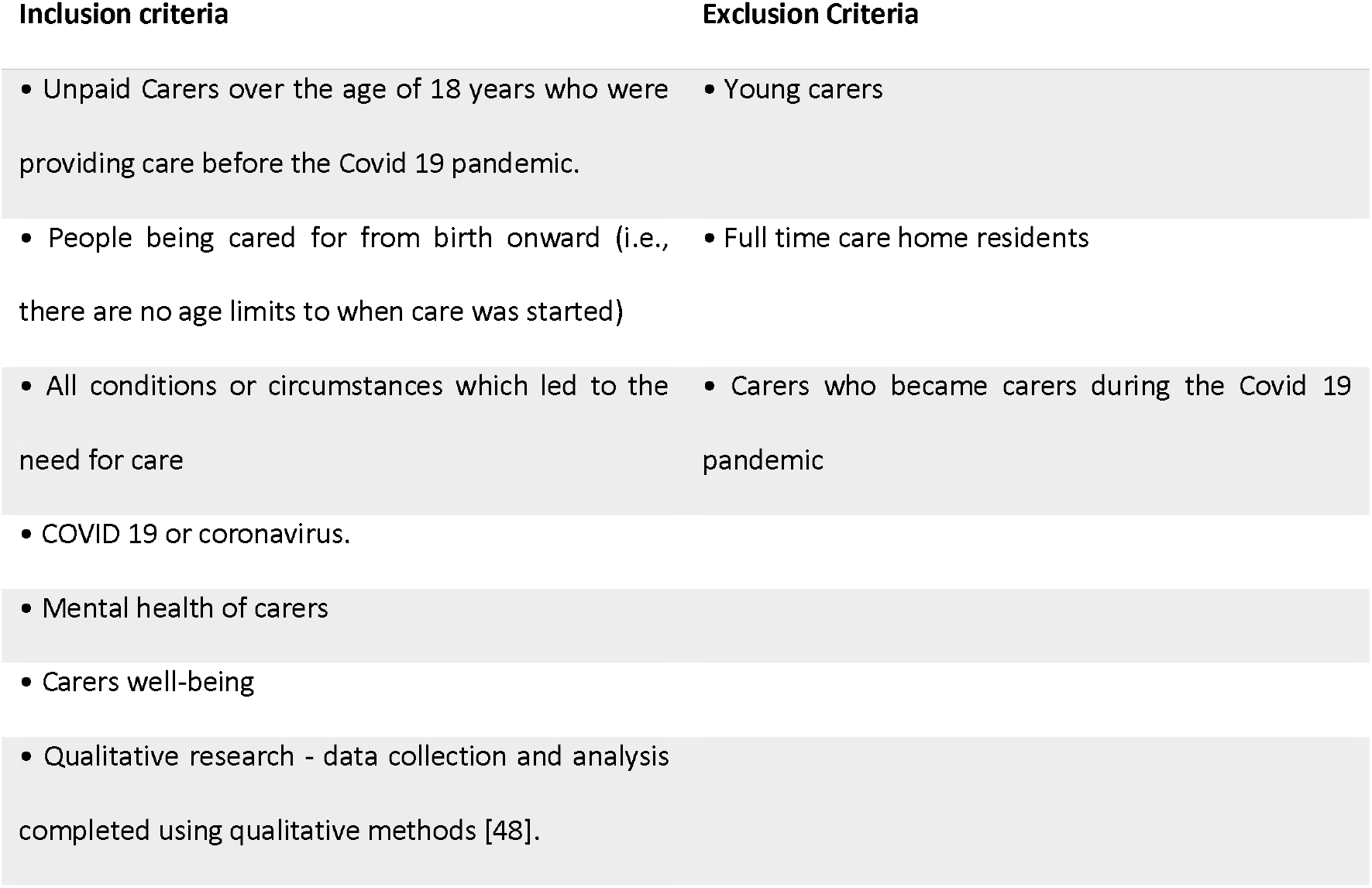
Inclusion and exclusion criteria

| Inclusion criteria | Exclusion Criteria |
| --- | --- |
| Unpaid Carers over the age of 18 years who were providing care before the Covid 19 pandemic.<br>People being cared for from birth onward (i.e., there are no age limits to when care was started) | Young carers<br>Full time care home residents |
| All conditions or circumstances which led to the need for care<br>COVID 19 or coronavirus. | Carers who became carers during the Covid 19 pandemic |
| Mental health of carers<br>Carers well-being |  |
| Qualitative research - data collection and analysis completed using qualitative methods [48]. |  |

P - Family or friend caregivers over the age of 18 who do not get paid to provide care.

I – The impacts the measures put in place to combat the COVID 19 pandemic have on the mental health and well-being of unpaid carers.

Co - Isolation, social distancing and shielding during the COVID 19 pandemic.

Citation searching and hand searching is recommended in case relevant studies have been overlooked in the database searches [45, 46, 50], this was not done because the articles selected for appraisal were very recent and had not been cited.

### 2.2 Data sources and search strategy

A range of databases were searched; CINAHL plus [51], Ovid (Embase, MEDLINE, APA PsycINFO) [52] and The Cochrane [53] library, to reduce the risk of excluding relevant research [45]. Google Scholar and more were searched for grey literature, a full list is available in the research protocol (Appendix A).

The scoping search identified key words and Medical Subject Heading (MeSH) terms to be used and were refined before the search began (Table 2). An exploration of MeSH terms in all three databases searched revealed the term ‘caregiver’ was the only term applicable to unpaid carers. However, it is also a term used to describe parents and health care workers in several fields and so key words with a mixture of Boolean operators, truncation and proximity searching were used in varying ways to conduct the search.

**Table 2.**
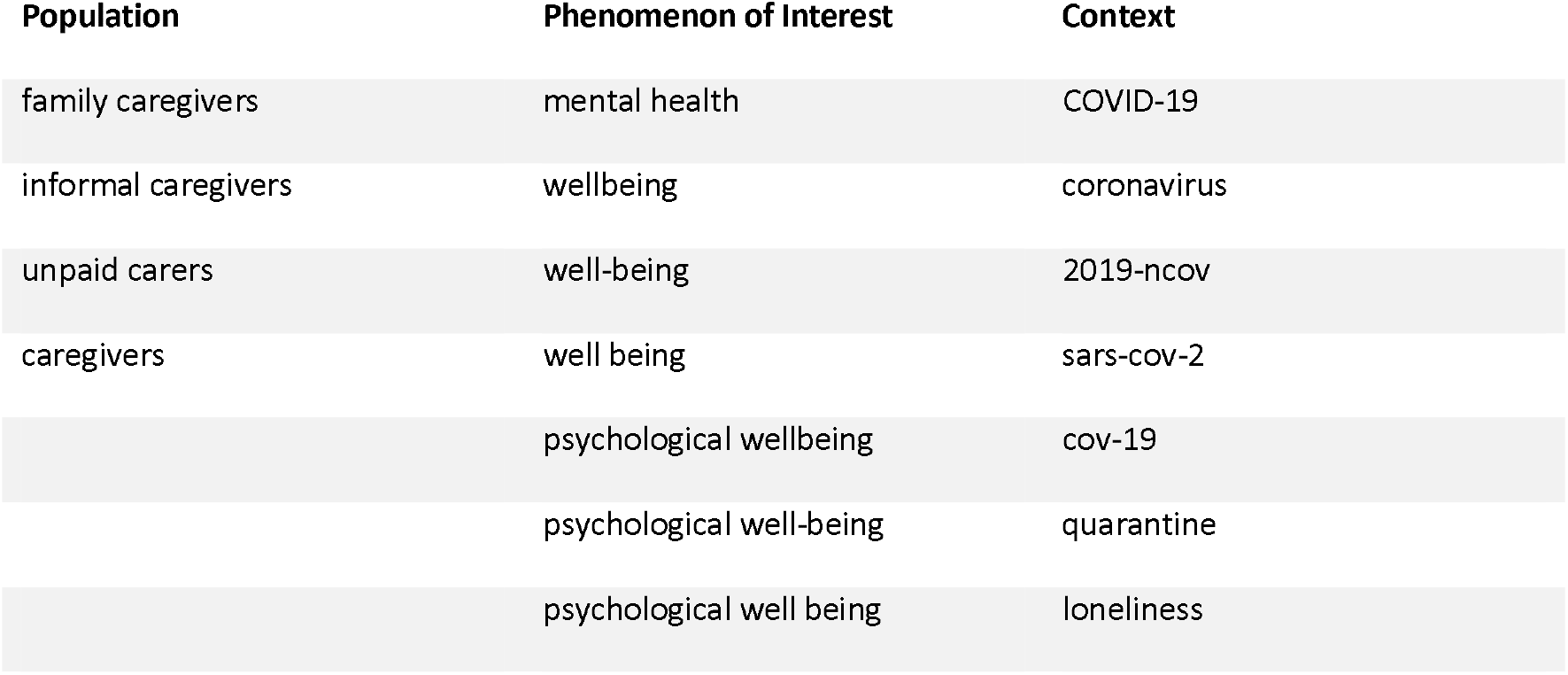
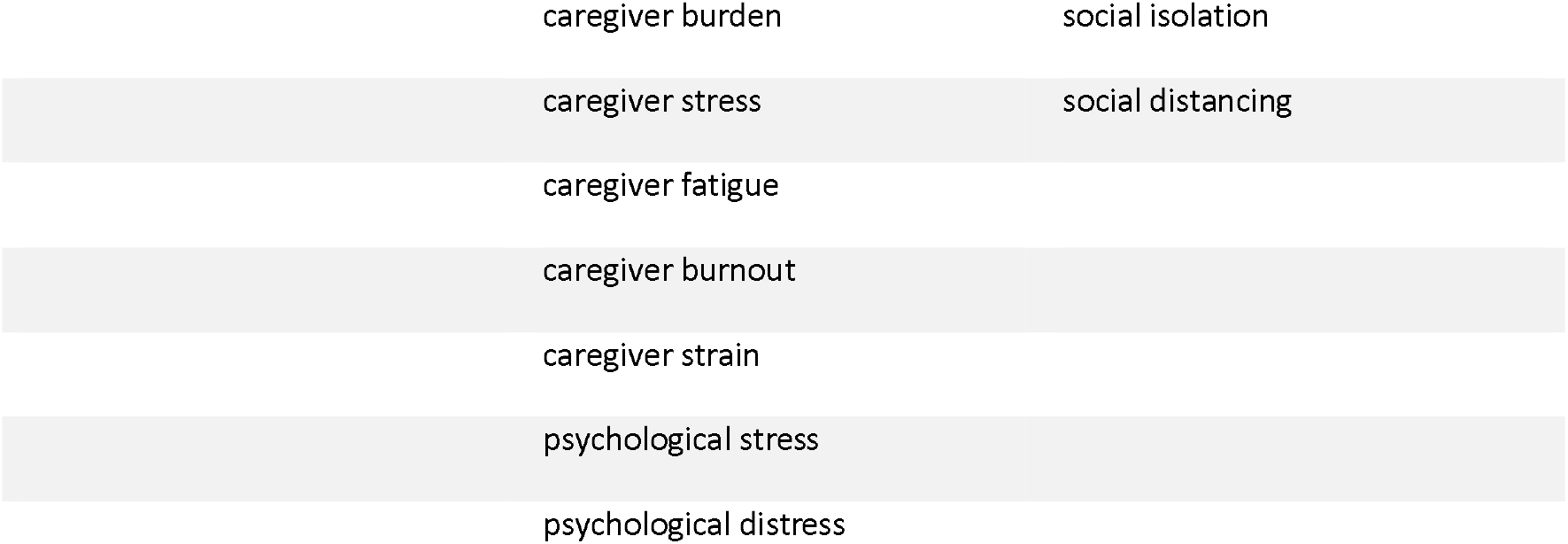
Keywords

No limits were placed on the keyword searches. The initial searches were carried out between the 18^th^ and 25^th^ of August 2020. See Table 3 for samples of the database search. The full database and grey literature searches are available in Appendices C and D.

**Table 3.**
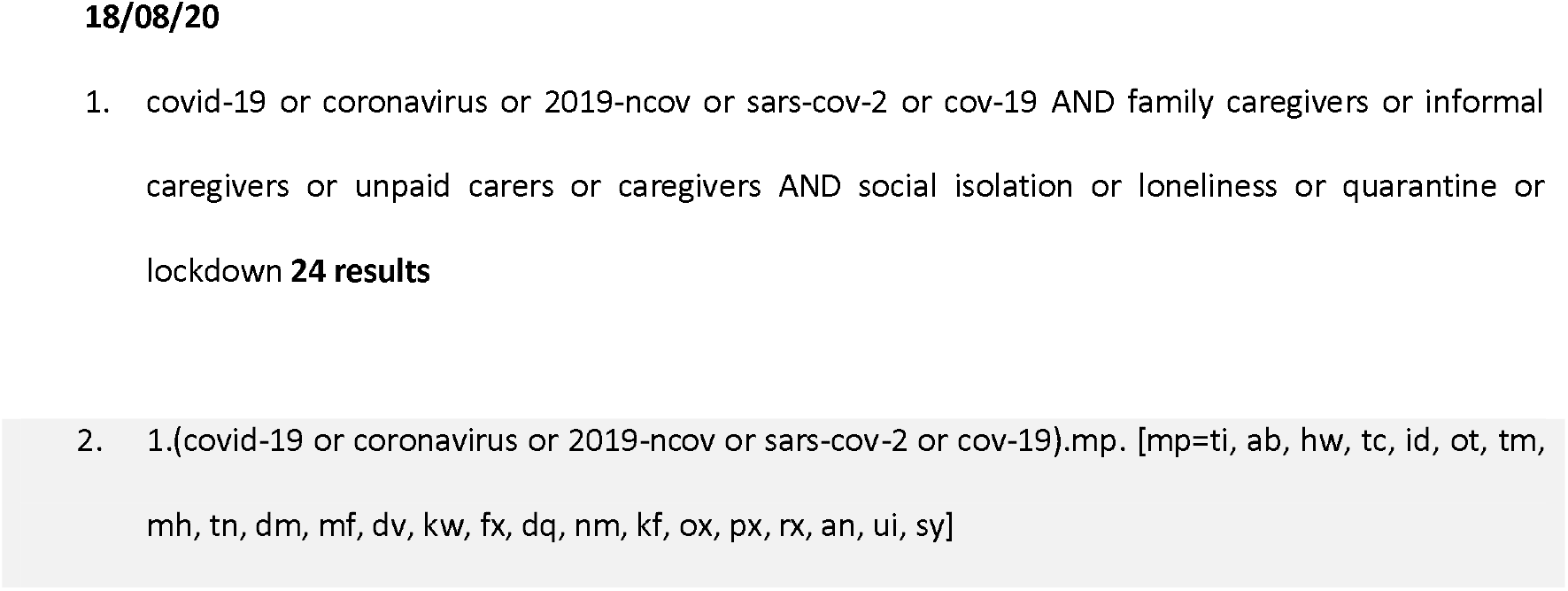
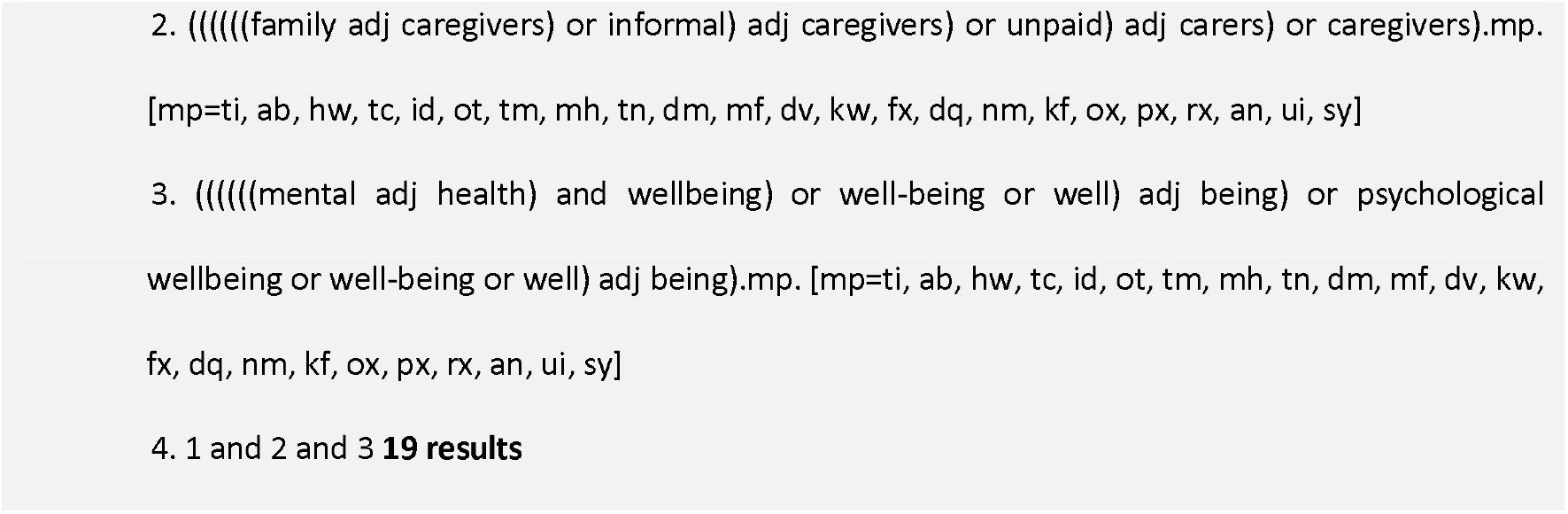
Sample of the CINAHL and Ovid search

After screening, two studies were selected for appraisal. As the COVID 19 pandemic is ongoing a repeat search was conducted between the 30^th^ of October and the 5^th^ of November.

Reference management software Endnote X9 [54] was used which enabled results from different databases to be combined and automatic removal of duplicate studies. A manual review of the studies also identified duplicated studies which were removed.

### 2.3 Screening methods

Screening was carried out in three stages by the author. First using titles, then reading abstracts and lastly reading full articles to determine eligibility for appraisal. A full list of excluded articles is available with reasons for exclusion (Appendix E), the inclusion and exclusion criteria were used to screen studies.

### 2.4 Rationale for appraisal

There are several viewpoints on the value of critically appraising primary qualitative research, principally because qualitative research has several methodologies and underlining theoretical approaches [43, 47, 55]. National Institute for Health and Care Excellence (NICE) [56] who use evidence-based research to develop health and social care guidelines and advice, list the CASP Qualitative checklist [57] as the preferred appraisal checklist for qualitative studies [58], as do the Cochrane policy group [48].

Therefore, the CASP [57] appraisal tool for qualitative research studies was used to assess the methodological strengths and limitations and to determine inclusion or exclusion in tandem with the data extraction tool (Appendix A).

## 3 Results

### 3.1 Study selection

Study selection was completed using the initial and repeat search (Figure 1). All excluded records are available in Appendix E and records excluded after assessment of eligibility in Appendix F. Between the two searches a total of 168 records were identified for screening. After duplicates were removed and articles excluded 33 full records were screened. Five records were selected for inclusion, two of those were excluded after appraisal.

**Figure 1.**
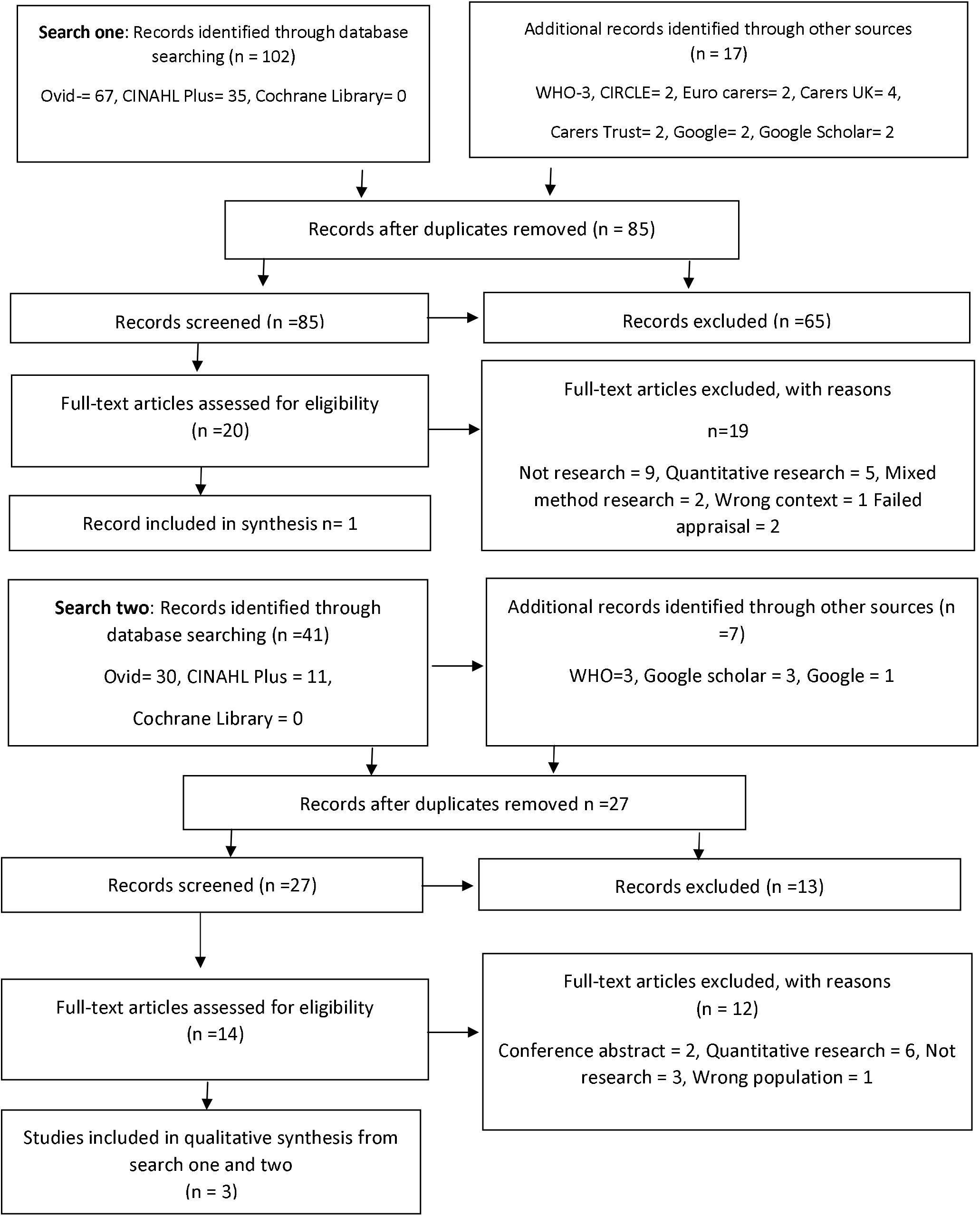
Adapted PRISMA Flowchart ^1^Adapted from: Moher, D., Liberati, A., Tetzlaff, J. and Altman, D. (2009). Preferred reporting items for systematic reviews and meta-analyses: the PRISMA statement BMJ, vol. 334, p. b2535. [Online]. Available at: https://www.bmj.com/content/339/bmj.b2535.full?view=long&pmid=19622551 (Accessed 5th November 2020)

The initial database search retrieved 102 titles and the grey literature search retrieved 17 records for screening. Once duplicates were removed and titles were excluded, 20 articles were screened for inclusion. The majority of these were excluded as the wrong study type and two were excluded after appraisal leaving one record for inclusion. The repeat search retrieved 41 records from the databases and 8 from the grey literature search. Thirteen full records were screened after duplicate and excluded records were removed. This search concluded with two further records to be included, so three records were identified for inclusion in total.

### 3.2 Appraisal

The CASP qualitative research tool [57] separates three broad themes; validity of the results, what the results are and transferability of the results into ten questions to critically appraise evidence (see Appendix G). CASP [57] do not recommend a scoring system, so an overall judgement was made after the appraisal using the terms; weak, for studies with several weaknesses and limitations, medium, for studies which had some weaknesses and limitations and strong for studies which had predominantly strengths. Five studies were appraised by the author (Table 4) and two were excluded.

**Table 4.** Summary of appraisals

| *Study* | *Dhavale et al. [37]* | *Vaitheswaran et al. [38]* | *Savla et al. [39]* | *Giebel et al., [40]* | *Clark-Bryan et al., [41]* |
| --- | --- | --- | --- | --- | --- |
| **1. Was there a clear statement of the aims of the research?** | yes | yes | yes | yes | yes |
| 2. Is qualitative methodology appropriate? | yes | yes | yes | yes | yes |
| 3. Was the research design appropriate to address the aims of the research? | can't Tell | yes | can't tell | can't tell | yes |
| 4. Was the recruitment strategy appropriate to the aims of the research? | can't Tell | yes | can't tell | yes | yes |
| 5. Was the data collected in a way that addressed the research issue? | can't tell | can't tell | can't tell | yes | yes |
| 6. Has the relationship between researcher and participants been adequately considered? | no | can't tell | no | yes | no |
| **7. Have ethical issues been taken into consideration?** | can't tell | can't tell | can't tell | yes | can't tell |
| **8. Was the data analysis sufficiently rigorous?** | no | can't tell | can't tell | yes | yes |
| **9. Is there a clear statement of findings?** | yes | yes | yes | yes | yes |
| **10. How valuable is the research?** | Weak | Medium | Weak | Strong | Strong |

There is also some debate about the exclusion of research results deemed as poor quality. These debates include the opinions that they should be excluded, researchers should use the audit trails of the research to determine their exclusion [47, 55] and only findings which cannot be supported by data should be excluded [55]. This research has used a data extraction tool alongside the appraisal process and so the two excluded studies were not only deemed to be of poor quality, they also did not provide sufficient information to extract data which was required to provide consistency and transparency for this review.

### 3.3 Data extraction

Use of a data extraction tool is recommended [48] to facilitate consistency of extracted data. One was developed using the NICE [58] template (in Appendix A). Data extraction has been summarised (Table 5) and a characteristics of included studies is provided in Appendix H.

**Table 5.** Summary of data extraction

|  |  |  |  |
| --- | --- | --- | --- |
| **Study** | ***Vaitheswaran et al. [38]*** | ***Giebel et al. [40]*** | ***Clark-Bryan et al. [41]*** |
| **Population** | Carers of people who have dementia. | Carers of people who have dementia. | Carers of people who have anorexia nervosa. |
|  | Female carer majority | Female carers majority<br>Spouses provided majority of care followed by adult children.<br>Mean age of carers is 60 years old. | Female carers majority<br>Parents<br>Mean age of carers is 54 years old. |
|  | Wives provided most care. |  |  |
|  | Mean age of carers is 54 years old. | Socioeconomic status included. |  |
|  | Socioeconomic status included. |  |  |
| **Interest** | Experience of carers of people who have dementia during the COVID 19 pandemic. | Impact of closures of social care support services due to COVID 19. | Effect of lockdown measures on patients and their carers. |
| **Context** | after the introduction of lockdown measures 24<sup>th</sup> of March in India. | after the introduction of lockdown measures on 23rd March in the UK. | after the introduction of lockdown measures on 23<sup>rd</sup> March in the UK. |
| **Results** | Inductive analysis with supportive quotations. | Inductive analysis with supportive quotations. | Inductive and deductive analysis with supportive quotations<br>clinical implications of their research |

### 3.4 Coding and synthesis

Coding was carried out by the author; codes were developed as quotations were selected and grouped from each study concluding in the development of analytical themes identified using thematic synthesis [59].

Research findings from the results, findings and discussion sections of the included studies were used. The studies were read in full again after appraisal and quotations (both direct and from the authors of the studies) were highlighted on paper copies of the studies. The studies were read again in electronic form and quotations were selected and input into the EPPI Reviewer4 software [60] with descriptive headings.

As one author completed the coding, a comparison between the highlighted quotations and the quotations in the software was made to ensure relevant information was not omitted. The quotations and themes were read through and assigned group headings (Appendix I). The quotations and descriptive themes were read again and four analytical themes emerged: immediate worries, adapting to change, post pandemic fears and use of technology (see Appendix J). Table 6 in section 3.4.2 contains quotations relevant to each theme.

**Table 6.** Quotations

| *Immediate worries* | *Adapting to change* | *Post pandemic fears* | *Use of technology* |
| --- | --- | --- | --- |
| **Health and/or behaviour deterioration** | **Routines and/or activities** | **Social care support** | **Access to services** |
| '[She's] got high levels of anxiety and OCD so you can imagine that, things are not easy.' (May 5, 2020) [41, p.831] | '...So he had a really quite an active social life really and I spent a lot of time just organising things so some of the, there was a lot that he could do. ....' Female carer (Spouse) to PLWD with young-onset dementia, Interview 13 [40, p. 4] | 'There was a general concern and worry about the uncertainty of the re-opening of social support service provision in the future and how this would be | 'The findings revealed that eating disorder services across the UK had reduced provision of face-to-face care and moved towards delivering remote support where possible, via phone calls or video conferencing.' (Authors) [41, p. 832] |
| 'I think that the longer this goes on the more he won't be able to go back to the things that he was doing.....That's another |  |  | 'I do not know how to use a computer or a big (smart) phone.' [38, p. 1191] |
|  |  |  | **Care delivery** |
| *thing because church has* |  | *achieved.* |  |
| *stopped, he used to go up* | *'I am not able to go to* | (Authors) [40, | *'Some services have adapted in* |
| *to church and he'd go and* | *work.. . . . so there has* | p.4] | *providing remote support in the form of* |
| *see my mums grave and* | *been a loss in income.'* |  | *Zoom internet meetings of previous peer* |
| *all of that's stopped as* | [38, p. 1189] |  | *support groups. Some day care centres* |
| *well.' Female carer* |  | *'With the* | *provided regular phone calls with the* |
| *(Daughter), Interview 44* | *"You just want some* | *additional* | *unpaid carers.'* (Authors) [40, p.5] |
| [40, p. 3]. | *time apart and going to* | *challenge of* |  |
|  | *work does that and you* | *when normal* | *'The sense of being "abandoned" by* |
| *'Isolation and* | *need to have that break* | *social support* | *services was exacerbated when promises* |
| *overprotection affects the* | *as a carer sometimes,* | *services are* | *of remote support were not delivered.'* |
| *quality of life of the* | *don't you?" (May 16,* | *resuming, many* | (Authors) (41, p.832) |
| *elderly, especially those* | 2020) [41, p. 831] | *carers are* |  |
| *with dementia. After all,* |  | *becoming even* | *'Moreover, not everyone has access to* |
| *what is the point in* | **Reduced physical** | *more* | *the Internet or knows how to use or can* |
| *protecting life if they* | **activity of people who** | *distressed.'*(Aut | *use Zoom,....' (Authors) [40, p.5]* |
| *cannot live it?' [38,* | **require care** | hors) [40, p. 6] |  |
| p.1188]. |  |  |  |
|  | *'My father cannot go out* | *'I'm just hoping* |  |
| **Protection from exposure** | *for walks here in the city,* | *all these* |  |
| **to COVID 19** | *which makes him very* | *services* |  |
|  | *upset and aggressive* | *eventually do* |  |
| *'We cannot explain to him* | *inside the apartment...'* | *come back and* |  |
| *about keeping a distance* | [38, p. 1190] | *they come back* |  |
| *from others and he will not* |  | *sooner rather* |  |
| *follow; he can become* | **Reduced support** | *than later* |  |
| *irritable on prompting'* |  | *because then* |  |
| [38, p.1187] | *'Suddenly losing control* | *and my Dad* |  |
|  | *over accessing support,* | *hasn't been out,* |  |
| *'Afraid to take her out.* | *which has often been* | *he hasn't been* |  |
| *Worry about her catching* | *difficult to obtain in the* | *the shops he* |  |
| *the infection.'* [38, p. 1189] | *first place before the* | *hasn't been* |  |
|  | *outbreak, was a frequent* | *obviously* |  |
| **Health of carer** | *worry.'* (Authors) [40, | *walking..... he* |  |
|  | *p.6]* | *really loves* |  |
| *'public health restrictions* |  | *being round* |  |
| *due to the virus outbreak* | **Increased carer** | *people.'* Female |  |
| *have led to increased carer* | **responsibility** | *carer* |  |
| *burden in many unpaid* |  | *(Daughter),* |  |
| *carers...' (Authors) [40, p.* | *Carers spoke about* | *Interview 24* |  |
| *6]* | *increased pressures and* | *[40, p. 5]* |  |
|  | *expectations on* |  |  |
| *'Don't know how my* | *themselves, particularly* |  |  |
| *husband will manage* | *as the primary carer.* |  |  |
| *with- out me.'* [38, p. | *(Authors) [41, p.831]* |  |  |
| *1189]* |  |  |  |

Full texts were re-read during the development of themes to ensure that the quotations were used within their original context to maintain the integrity of the primary research.

Frequency charts (Appendix K) display the number of studies which contributed to each sub-theme and were developed using EPPI reviewer 4 software [60]. Additionally, the summary of characteristics also provided a comparison of the studies.

#### 3.4.1 Theme 1 - Immediate worries/fears

##### Health and/or behaviour deterioration

Carers noticed that symptoms of existing conditions and illnesses had increased in the people they cared for after lockdown began and expressed concerns that their conditions would deteriorate. This was attributed to early hospital discharges, anxiety about COVID19, reduced support from the hospital [41] and closure of facilities and a lack of mental stimulation [40]. However, one carer questioned the value of the lockdown measures as she worried that they were reducing her mother’s quality of life through loss of social connections and activities [38].

##### Protection from exposure to COVID 19

Many carers believed the people they cared for did not have the capacity to understand the rules and so would not socially distance from other people [38, 40]. To address this, carers reduced the amount of time the people they cared for spent outside of their home. In some cases, these restrictions became a source of stress between the carers and the people they care for [40]. Other carers were afraid that the people they cared for would go out and contract COVID 19 [38].

##### Health of carer

One study [40] directly addressed the impact of increased dependency on carers; emphasising that paid carers are usually employed when they are needed. Despite this, in all three included studies [38, 40, 41] carers expressed more concern about the people they cared for and the impact of the restrictions on them.

#### 3.4.2 Theme 2: Adapting to change

##### Routines and/or activities

This was one of the most frequently occurring sub themes across all three studies. It is evident that the routines of both carers and the people they cared for were affected by the lockdown measures in many ways.

Numerous people who have dementia and had access to activities could no longer attend them because of the restrictions during lockdown. This caused concern to their carers as they were worried about how they could provide mental stimulation and activities for them to do at home [38, 40]. Routine was also expressed as playing an important role as a coping mechanism and recovery tool for people who had anorexia nervosa [41]. These changes meant that carers were providing more care than they had previously as they became responsible for filling the gaps in caused by the loss of access to social and physical activities.

Some carers had to stop work or work from home which resulted in a loss of income and difficulty in working effectively [38] and loss of respite for those who viewed time at work as respite [41]. Although one carer used it as an opportunity to reflect on their life and reconsider priorities which they saw as a positive opportunity [41].

##### Reduced physical activity of people who require care

Carers were also worried about lockdown restrictions starting to impact on the levels of physical activity of the people they cared for as they were unable to maintain the routines they had developed to keep them physically fit and active [38, 40].

##### Reduced support

This is interlinked with health and/or behaviour deterioration and changes to routines and activities and further demonstrates the knock-on effect of the lockdown measures on both the carers and the people they cared for which created new problems for carers to address; especially when many have had challenges with receiving or accessing respite to begin with [40].

##### Increased carer responsibility

During the lockdown period some carers felt overwhelmed by having to maintain daily household chores, care for their relatives and look after their and other family members well-being [41].

#### 3.4.3 Theme 3: Post pandemic fears

##### Social care support

Carers were thinking about how and when social support services would open and again expressed concern about the people they cared for and the impact of them not having access to their usual activities and routines [40], rather than any impact this may have also had on themselves.

Adding to the above concerns was the worry that social support services previously offered would no longer be suitable and what would happen if this was the case.

#### 3.4.4 Theme 4: Use of technology

##### Access to services

During the lockdown period more services were accessed via telephone calls and video conference calls [40, 41]. Several carers in a study carried out in India expressed that they would also like to have access to services in the same manner [38].

However, several carers also reported barriers to accessing services via video conference calls from low digital literacy to resistance from the person they cared for [38].

##### Care delivery

Some adjustments were made to deliver care via phone or video calls [40, 41]. One carer expressed improvements in care delivery because of more communication from the hospital and reassurance of physical support if required [41].

Some carers felt let down when they did not receive the support which was promised to them [41] and where support was adapted (in a few services) to be delivered through video calls, carers felt it did not fulfil the social needs of the people they care for [40]. And lastly, the issue of digital literacy, access to the internet and difficulty in the person being cared for using technology was raised again.

## 4 Discussion

A total of 112 studies were screened in an initial and an updated search. Of these, three studies were selected for inclusion, one which was conducted in India. Although the geographical context of this review was aimed at England, the study conducted in India was included as it met the inclusion criteria set for the review.

All included studies [38, 40, 41] discussed the experiences of carers during the first COVID 19 lockdown period. Two studies discussed social distancing [38, 40] and one [41] discussed shielding. The mental health and well-being of carers is discussed in all three studies. Although very few carers explicitly spoke about the impact of the lockdown measures on their mental health and well-being, the impacts were implicitly present and became apparent as the thematic synthesis of the results progressed.

To the author’s knowledge, this is the first systematic review on this topic and at the time it was conducted there was a paucity of relevant qualitative research. As such, this review has identified gaps in research and made suggestions for further research.

### 4.1 Interpretation of results

The earlier definition of mental health and well-being described what can affect mental health and well-being, specifically in relation to carers and included many factors. The results of this review reflect several of these factors, which impacted on the mental health and well-being of carers, resulting from the measures implemented during lockdown.

#### Themes 1, 2 and 3

It became evident across the included studies [38, 40, 41] and in themes 1, 2 and 3 that when given the opportunity to share their experiences, carers expressed more concern for the people they care for than they did their own needs. Previous studies have already identified the impact becoming a carer can have on the mental health and well-being of carers [16, 21, 22, 23, 24]. In particular, the care that is received by the individual they care for, may not only determine the level of practical burden experienced by the unpaid carer, but may also determine the individual’s day-to-day experiences and general well-being. This may create a further burden of increased concern in the unpaid carers [6], as reflected within the findings of this review. However, there is a lack of qualitative research carried out to explore the experiences of carers and identify or address their needs, as they specifically arise in relation to the pandemic.

Many carers of people who have dementia expressed concern about the withdrawal of support services and activities having a detrimental effect on the people they care for. During the first lockdown a report revealed that 82% of people surveyed who have dementia reported an increase in their symptoms, confirming the fears expressed by the carers in this review [61]. The same report showed that 95% of carers reported a detrimental effect from their increased caring responsibilities; ranging from exhaustion to injury form their caring role [61]. On a positive note, a study concluded that video conference calling between healthcare professionals, service users and their carers was found to be beneficial to people with cognitive impairment [61].

The studies in this review were focused on carers of people who have dementia [38, 40] and carers of adults diagnosed with anorexia nervosa [41] – specific conditions. Although carers expressed some similar concerns and experiences this may not be the case for other carers. Therefore, purposefully sampled research to explore the experiences of specific groups of carers may provide more detailed information of the impacts and would therefore be more useful.

After the first lockdown, the Department of Health and Social Care (DHSC) [62] published the COVID-19: Our Action Plan for Adult Social Care, a small section of this focused on the support of carers. Following this, a Carers Advisory Group was established, one of their roles was to provide advice on the DHSC [62] care plan [63]. A comprehensive list of recommendations has been put forward to the DHSC which focuses on improving the health and well-being of carers and maintaining carers’ ability to care [63]. Given how relevant this is to the focus of this review, future research evaluating the implementation of these recommendations would be valuable.

The measures implemented during the first lockdown have led to an increased reliance on carers to maintain the health, safety and well-being of the people who they care for, which has already been recognised as a risk to the health and well-being of carers during the current pandemic [34]. The review results support survey findings, which revealed that carers’ roles had increased because of discontinued and/or reduced support services [16, 31] most of which had not been fully resumed after the first lockdown [32]. The results of this review also add to existing quantitative research [64, 65, 66, 67] and voices of concern [21, 61, 68], which point to the likelihood of carer burnout during the current pandemic.

Positively, recently published research demonstrated the resilience and resourcefulness of carers in their study who had adapted to the new conditions [69]. Carers used self-care strategies, humour and peer support to maintain their well-being partly facilitated through digital peer support [69].

#### Theme 4

Generally, carers were interested in the use of phone calls or video calls to maintain contact with services. Difficulties in the person requiring care using technology or using the devices needed to access technology, access to the internet and low digital literacy were raised as barriers to accessing services using phone and/or video calls.

These difficulties were also identified in a recent report [70], although these barriers were removed with support such as in accessing devices, training and more. Use of technology was found to be beneficial for the well-being of both carers and the people they cared for [70].

Increased use of technology to deliver and receive care is already a priority as part of the NHS Long Term Plan [71]. Furthermore, research on the benefits of using various digital resources to improve the health and well-being of carers and the people they care for during the pandemic is emerging. For example, research showed that helping people to connect using video calling to access peer support was proven to be beneficial to both carers and people who have dementia [69]. Additionally, Essex, Kent and Suffolk county councils identified a useful device which was trialled with some of their service users and their carers [72]. The device had several uses including the facility to use video calling between the user and their health care team [72].

A carer who called into a radio programme raised the point that for the first time since she became a carer she felt included in the world around her during the pandemic [35]. People started online activities and groups which she could not be a part of when she became a carer, because she could not leave the house as often as she once could [35]. There is some evidence that digital technology in various forms is having a positive impact on carers and the people they care for during the pandemic. However, this has been mostly from the perspective of maintaining links between healthcare services and users or focussed on users who have dementia and their carers. Therefore, research on the use of digital platforms to facilitate social activities for carers as well as research on digital literacy and access to digital technology among carers could be beneficial during and after the COVID 19 pandemic.

On the 5^th^ of January 2021 England started its’ third lockdown period [73] and although there are now approved vaccines being rolled out [74], it is still unknown when life will return to what it was before the pandemic. So, it is important to ensure that carers are supported to continue their roles with as little detrimental impact on their own mental health and well-being as possible.

## 5 Limitations

This review has several limitations. A single author has completed this review as part of a master’s degree; having more than one reviewer involved in each stage of a systematic review ensures that individual subjectivity does not affect the interpretation and results of the review [75]. Therefore, the research may be biased to the author’s subjective interpretation of the studies selected and reviewed.

The review was carried out after the first lockdown during the COVID 19 pandemic which is ongoing and there was limited information available on the topic at the time. It is expected that there is new research emerging as the pandemic continues.

The included studies focused on carers of people who were diagnosed with two conditions. Consequently, it is possible the results of the review are biased towards the experiences of carers who participated in the included studies.

Many studies were excluded due to the methodology of the research because this review was intended to be a review of qualitative research. Several surveys were excluded as quantitative research which could have contributed to the analysis of the review topic. Therefore, a review of both qualitative and quantitative research may have provided a richer insight into the review topic.

And lastly limiting the research to studies reported in English may have omitted relevant research. COVID 19 is a pandemic so there may be relevant research from countries that did not report their research in English.

## 6 Recommendations

1. Purposeful sample research to determine the needs of groups of carers during the COVID 19 pandemic.
2. Future research on the implementation of the Carers Advisory Group [63] recommendations to the DHSC.
3. Research on digital literacy and access to digital technology among carers.
4. Research on the use of digital platforms to facilitate social activities for carers during and after the COVID 19 pandemic.

## 7 Conclusion

The lockdown measures put in place to address the COVID 19 pandemic have resulted in increased reliance on carers and put them even more at risk of carer burnout. Carers express more concern for the health and well-being of the people they care for than their own health and well-being and more support is needed for carers to be able to sustain their caring role and reduce the detrimental impacts of increased reliance on them. Carers could benefit from accessing support from both health and social care services and peer support using technology such as video conference calling or online activities.

## Data Availability

All data is available in this paper and in the supplementary materials.

## Prior presentation(s)

None to declare.

## Supplementary materials

Appendices A-K

CAS: Critical Appraisal Skills Programme
DHSC: Department of Health and Social Care
ENTRE: Enhancing Transparency in Reporting the synthesis of Qualitative Research
MeSH: Medical Subject Heading
NICE: National Institute for Health and Care Excellence
NIH: National Institute for Health Research
PICo: Population, phenomenon of Interest and Context
WHO: World Health Organisation

