## Appendices for "The impact on the mental health and well-being of unpaid carers affected by social distancing, self-isolation and shielding during the COVID 19 pandemic in England – a systematic review": Appendices A-K.docx

Research Protocol

**Formulation of the review question.**

The question to be addressed in this review has been formulated using PICo as follows:

P- Family or friend caregivers over the age of 18 who do not get paid to provide care.

I – The impact the measures put in place to combat the COVID 19 pandemic have on the mental health and well-being of unpaid carers.

Co - isolation, social distancing and shielding during the COVID 19 pandemic.

Qualitative research will be systematically reviewed to explore the experience and views of carers as well as what facilitates positive mental health and well-being for unpaid carers during the current pandemic. The question which will be asked to address the research question is:

• How is the mental health and well-being of unpaid carers in England affected by the current measures in place to reduce the spread of the COVID 19 pandemic?

**Condition or domain being studied.**

On the 11th of March 2020, the WHO declared a coronavirus - COVID 19 Pandemic (WHO 2020a). This prompted the following HM Government (2020) measures: staying at home and keeping a distance from other people (known as social distancing), the closure of certain businesses and venues and gatherings of more than two people were stopped. Clinically vulnerable groups people over 70 years old and all people who are invited to have a yearly Flu vaccine due to their health conditions (HM Government 2020) were identified. The concept of shielding was also introduced for people who are clinically extremely vulnerable who needed to self-isolate and take extra precaution to protect themselves from the virus (PHE 2020).

In response to the COVID19 pandemic Carers UK and Carers Trust (2020) released a joint statement in which they emphasise an increased risk of loneliness and isolation for carers. Following this, research carried out by Carers UK (2020) revealed that burnout is a concern of 55% of the carers, the COVID19 outbreak has caused 70% of carers to increase the care they provide and the closure or reduction of local services has made 35% of carers provide more care.

**Context.**

Using figures from 2014, the ONS (2016) estimate that unpaid adult carers provided care which would have cost £56.9 billion if paid carers or nursing assistants were employed - an increase of 45.8% from 2005. This figure is expected to continue increasing due to an aging population.

The Family Resources Survey 2018/2019 revealed that 7% of the survey respondents were providing unpaid care: of these 57% were providing care for someone who does not live with them and 45% were caring for someone living in the same household (DWP) 2020). Unpaid carers are being increasingly recognised and valued for the contribution they make to society.

COVID 19 is an infectious disease caused by a newly discovered coronavirus which has no vaccine or treatment, it is spread from an infected person to another person through coughing or sneezing; expelling droplets of saliva or discharge from the nose (WHO 2020b and CDC 2020). Measures to reduce its spread include but are not limited to avoiding close contact with other people, self-isolating if you are unwell and frequent hand washing for a minimum of 20 seconds (WHO 2020c and CDC 2020).

**Main outcomes**

1. An exploration of the effect of caring on carers mental health and well-being

2. Contribution to an increasing evidence base of the impact of the lockdown, social distancing, self-isolation and shielding on carers, through a synthesis of existing qualitative information.

3. Identification of measures which facilitate good mental health and well-being during the COVID 19 Pandemic which can be used in other contexts. Such as in future epidemics or unexpected situations.

**Participants/population.**

The word carer will be used throughout this research to refer to adults (18 years and above) who are providing care to a relative, neighbour or friend without being paid (Department of Health and Social Care (DHSC) 2018). Care can be provided in many ways including help with household chores or help with eating meals (Department for Work and Pensions (DWP) 2020). Help may be needed due to mental health problems, illness, disability, frailty or substance misuse problems (DHSC 2018).

Inclusion criteria:

• Unpaid Carers over the age of 18 years who were providing care before the Covid 19 pandemic

• People being cared for from Birth onward (i.e., who require care at any age)

• All conditions or circumstances which led to the need for care

• COVID 19 or coronavirus.

- Mental health of carers
- Carers well-being

Exclusion Criteria:

• Young carers

• Full time care home residents

• Carers who became carers during the Covid 19 pandemic

**Exposures**

Physical and emotional exhaustion and mental health problems can be experienced by carers (Parkinson et al. 2017 and Carers Trust 2015) and quality of life can be affected negatively by taking on a caring role (Galiatsatos et al. 2017 and Parkinson et al. 2017). A survey carried out by Carers UK (2019) showed that 45% of carers expected a reduction in their quality of life after taking on a caring role. Furthermore, the NHS England (2019) NHS Long Term Plan acknowledges that carers are twice as likely to experience poor health, in part due to stress and social isolation: in comparison to the general population.

In May 2020, the UK Governments DCMS and OCS (2020) launched a Loneliness Covid-19 Grant Fund aimed at reducing the impact of self-isolating and social distancing, especially for people who are considered to be vulnerable. Some of this has been awarded to the Carers Trust (Howells 2020). CIRCLE (2020) provides some evidence, that the mental well-being of carers has been negatively impacted by the current COVID 19 pandemic.

The following will be explored in relation to the impact on and experience of carers during the Pandemic.

Interests:

Mental health

Social isolation

Well-being

Burnout

Lockdown

Self-isolating

Shielding

Quality of life

**Types of study to be included.**

This review aims to explore the experiences and perceptions of carers and will therefore use qualitative information from sources that report the experiences and views of carers.

Mixed method and quantitative studies will be excluded.

**Searches.**

Database searching using CINAHL plus (2020), Ovid (Embase, MEDLINE, APA PsycINFO) (2020) and Cochrane (2020).

A hand search of the University of Manchester Library using the predetermined search words.

Searches from October 2019, worldwide and in English.

Initial MeSH headings: Caregivers, social isolation, stress; psychological, burnout; psychological.

Initial key word search terms: COVID 19, family carers, family caregivers, informal carers, unpaid carers, lockdown, social distancing, self-isolating and shielding.

Citation searching of both database search results and grey literature.

Grey Literature online – such as government, Health and Social Care and third sector websites in England and worldwide will be used to obtain evidence, listed below:

WHO

Rethink

Public Health England

Centers for Disease Control and prevention

MIND,

CIRCLE

NHS England

HM Government

Department for Work and Pensions

Department for Digital, Culture, Media & Sport and Office for Civil Society

The Health Foundation

Carers UK

Carers Trust and more

Hand search internet search terms will include key terms such as ‘carers and social distancing’ ‘carers and Covid 19’ ‘Covid 19 pandemic and mental health’, ‘Health and well-being during COVID 19’.

Authors will be contacted to request further information where required.

**Data extraction (selection and coding).**

Tabo Akafekwa (TA) will use the inclusion and exclusion criteria to select eligible papers. Abstracts will be screened to determine suitability; full articles will be read initially where abstracts do not provide sufficient information. Eligible articles will be read in full to assess eligibility. The Moher et al. (2009) PRISMA flow diagram will be used to present the research process and reference management software: Endnote (Web of Science Group 2020) will be used.

The CASP (2018) critical appraisal tool checklist for qualitative research will be used to assess the trustworthiness of the research evidence where appropriate.

Data extraction will be done using a pre-existing template (NICE 2018) (see appendix 1) and coding will be done using EPPI- reviewer 4 software (Thomas et al., 2010) and will consist of information from research findings and results sections.

TA will receive advice and support from dissertation supervisor Professor Arpana Verna (AV)

**Risk of bias (quality) assessment.**

Characteristics of studies to be assessed:

Rationale for the research

Participants

Setting of the research

Methodology

Results/findings

Discussion

Strengths and limitations

The Tong et al. (2012) ENTREQ statement will be used as a guideline for the review. Additionally, a summary of completed CASP (2018) qualitative checklists and AACODS Checklist (Tyndall 2010) for grey literature will be made available.

**Strategy for data synthesis.**

Key themes will be identified using thematic synthesis (Thomas and Harden 2008). This will be done by TA through inputting research findings/results using EPPI- reviewer 4 software (Thomas et al., 2010). Initially this will be done line by line identified with codes made by TA: initial codes will consist of descriptions of the text. This will continue until all data has been coded. Initial codes will then be reviewed and grouped as the research progresses resulting in fewer codes and emerging themes which will then be discussed. Thematic synthesis has been chosen as a method because secondary information (including quotations from results and findings sections of research papers) will be interpreted and combined culminating in new themes or theory.

This will be carried out with the advice and support of AV.

**Conflicts of interest.**

The reviewer has a professional history with unpaid carers as a former RSCN and has personal experience of being a carer.

Appendix 1

Data extraction template

| Bibliographic reference: authors, year, article title, journal, volume, pages. |  |
| --- | --- |
| Research question: what were the research questions?  Theoretical approach: what theoretical approach (for example, grounded theory, interpretive phenomenological analysis) does the study take (if specified)? |  |
| Data collection: how were the data collected? Give details of:  methods  by whom  when. |  |
| Method and process of analysis: what methods were used to analyse the data (for example, constant comparative method)? |  |
| Population and sample collection: what population was the sample recruited from? Include the following information:  - how they were recruited (for example, specify the type of purposive sampling)  -how many participants were recruited  - specific exclusion criteria  - specific inclusion criteria. |  |
| Settings: The settings where the qualitative study was undertaken. |  |
| Results: Key themes: list all relevant to this review (with illustrative quotes if available). |  |
| Source of funding: for example, the Department of Health or Economic and Social Research Council, and the role of funding organisations. |  |
| Quality assessment: Document any concerns about quality which can be used to provide an overall assessment of each study (e.g., rating from quality  checklist) for use in CERQual assessment |  |
| Limitations: both those identified by the authors and those identified by the reviewer. | Authors:  Reviewer: |

**Appendix B - ENTREQ statement**

| No | Item | Guide and description | Page number |
| --- | --- | --- | --- |
| 1 | Aim | To explore the impact on the mental health and well-being of unpaid carers affected by social distancing, self-isolation and shielding during the COVID 19 pandemic in England | 11 |
| 2 | Synthesis methodology | Thematic synthesis | 23 |
| 3 | Approach to searching | Pre-planned and comprehensive | 11-13 |
| 4 | Inclusion criteria | Inclusion criteria:  • Unpaid Carers over the age of 18 years who were providing care before the Covid 19 pandemic  • People being cared for from Birth onward (i.e. who require care at any age)  • All conditions or circumstances which led to the need for care  • COVID 19 or coronavirus. | 12 |
| 5 | Data sources | CINHAL Plus, Embase, APA PsychInfo, Ovid MEDLINE and Cochrane Library via Evidence Based Medicine Reviews (EBMR) via Ovid online. As well as pre-selected websites for grey literature listed in the research protocol Appendix 1 | 13, 60 |
| 6 | Electronic Search strategy | Full search strategy documented in Appendices 4 and 5 | 14 – 18, 83, 88 |
| 7 | Study screening methods | Screening and study selection were carried out by one reviewer. Search results were screened using titles and citations were added to EndNoteX9 reference management for further screening. Abstracts were read to select articles to be read in full. | 18 |
| 8 | Study characteristics | Available in Table 7 | 29 |
| 9 | Study selection results | Results are documented in Figure. 1, excluded articles are documented in Appendix 7 | 25 and 95 |
| 10 | Rationale for appraisal | Studies were appraised for methodological strengths and limitations. Medium and strong items were included in the review. | 23-24 |
| 11 | Appraisal items | The CASP (2018) tool for qualitative research was used. | 27 |
| 12 | Appraisal process | One person carried out appraisal of studies as part of a student dissertation project |  |
| 13 | Appraisal results | A summary is provided in Table 6. Full appraisals available in Appendix 8 | 27, 98-104 |
| 14 | Data extraction | Data was extracted manually using a template from NICE (2018). Data was extracted from the results, discussion and conclusion sections of primary studies. Template available in Appendix 6 | 92 |
| 15 | Software | EPPI Reviewer 4 (Thomas et al., 2010) |  |
| 16 | Number of reviewers | One person carried out appraisal of studies as part of a student dissertation project |  |
| 17 | Coding | Line by line coding was done in 3 stages: selection, development of descriptive themes and development of analytical themes | 32-33 |
| 18 | Study comparison | Comparisons were made during each coding stage and a frequency pie chart was made to visually represent how many studies contributed to each theme. Frequency pie charts available in Appendix 9 | 33, 104 |
| 19 | Derivation of themes | Inductive coding was carried out | 32-33 |
| 20 | Quotations | Quotations are provided in the results sections of the study for each analytical theme | 34-43 |
| 21 | Synthesis output | The discussion section provides a brief summary of the results and concludes the analysis of the review results. Recommendations are made for future research | 36, 44-48 |

**Appendix C - Database searches**

**Results in bold**

CINHAL Plus

**18/08/20**

Embase to August 17, 2020, APA PsychInfo and Ovid MEDLINE(R) and Epub Ahead of Print, In-Process & Other Non-Indexed Citations, Daily and Versions(R)1946 to August 17, 2020

Titles and key words scanned along with titles – no abstracts read.

**18/08/20**

1. 1.(covid-19 or coronavirus or 2019-ncov or sars-cov-2 or cov-19).mp. [mp=ti, ab, hw, tc, id, ot, tm, mh, tn, dm, mf, dv, kw, fx, dq, nm, kf, ox, px, rx, an, ui, sy]

2. ((((((family adj caregivers) or informal) adj caregivers) or unpaid) adj carers) or caregivers).mp. [mp=ti, ab, hw, tc, id, ot, tm, mh, tn, dm, mf, dv, kw, fx, dq, nm, kf, ox, px, rx, an, ui, sy]

3. ((social adj isolation) or loneliness or quarantine or lockdown).mp. [mp=ti, ab, hw, tc, id, ot, tm, mh, tn, dm, mf, dv, kw, fx, dq, nm, kf, ox, px, rx, an, ui, sy]

4. (social adj distanc*).mp. [mp=ti, ab, hw, tc, id, ot, tm, mh, tn, dm, mf, dv, kw, fx, dq, nm, kf, ox, px, rx, an, ui, sy]

5. 3 or 4

6. 1 and 2 and 5 **91**

2. 1.(covid-19 or coronavirus or 2019-ncov or sars-cov-2 or cov-19).mp. [mp=ti, ab, hw, tc, id, ot, tm, mh, tn, dm, mf, dv, kw, fx, dq, nm, kf, ox, px, rx, an, ui, sy]

2. ((((((family adj caregivers) or informal) adj caregivers) or unpaid) adj carers) or caregivers).mp. [mp=ti, ab, hw, tc, id, ot, tm, mh, tn, dm, mf, dv, kw, fx, dq, nm, kf, ox, px, rx, an, ui, sy]

3. ((((((mental adj health) and wellbeing) or well-being or well) adj being) or psychological wellbeing or well-being or well) adj being).mp. [mp=ti, ab, hw, tc, id, ot, tm, mh, tn, dm, mf, dv, kw, fx, dq, nm, kf, ox, px, rx, an, ui, sy]

4. 1 and 2 and 3 **19**

3. 1. (covid-19 or coronavirus or 2019-ncov or sars-cov-2 or cov-19).mp. [mp=ti, ab, hw, tc, id, ot, tm, mh, tn, dm, mf, dv, kw, fx, dq, nm, kf, ox, px, rx, an, ui, sy]

2. ((((((family adj caregivers) or informal) adj caregivers) or unpaid) adj carers) or caregivers).mp. [mp=ti, ab, hw, tc, id, ot, tm, mh, tn, dm, mf, dv, kw, fx, dq, nm, kf, ox, px, rx, an, ui, sy]

3. (((((((((caregiver adj burden) or caregiver) adj stress) or caregiver) adj fatigue) or caregiver) adj burnout) or caregiver) adj strain).mp. [mp=ti, ab, hw, tc, id, ot, tm, mh, tn, dm, mf, dv, kw, fx, dq, nm, kf, ox, px, rx, an, ui, sy]

4. (((psychological adj stress) or psychological) adj distress).mp. [mp=ti, ab, hw, tc, id, ot, tm, mh, tn, dm, mf, dv, kw, fx, dq, nm, kf, ox, px, rx, an, ui, sy]

5. 3 or 4

6. 1 and 2 and 5 **12**

4. 1. (covid-19 or coronavirus or 2019-ncov or sars-cov-2 or cov-19).mp. [mp=ti, ab, hw, tc, id, ot, tm, mh, tn, dm, mf, dv, kw, fx, dq, nm, kf, ox, px, rx, an, ui, sy]

2. ((((((family adj caregivers) or informal) adj caregivers) or unpaid) adj carers) or caregivers).mp. [mp=ti, ab, hw, tc, id, ot, tm, mh, tn, dm, mf, dv, kw, fx, dq, nm, kf, ox, px, rx, an, ui, sy]

3. ((mental adj health) or lonl*).mp. [mp=ti, ab, hw, tc, id, ot, tm, mh, tn, dm, mf, dv, kw, fx, dq, nm, kf, ox, px, rx, an, ui, sy]

4. 1 and 2 and 3 **47**

Cochrane Library via Evidence Based Medicine Reviews (EBMR) (Ovid Online)

**19/08/20**

1. 1. (covid-19 or coronavirus or 2019-ncov or sars-cov-2 or cov-19).mp. [mp=ti, ot, ab, tx, kw, ct, sh, hw]

2. ((((((family adj caregivers) or informal) adj caregivers) or unpaid) adj carers) or caregivers).mp. [mp=ti, ot, ab, tx, kw, ct, sh, hw]

3. ((((((mental adj health) and wellbeing) or well-being or well) adj being) or psychological) adj wel*).mp. [mp=ti, ot, ab, tx, kw, ct, sh, hw]

4. 1 and 2 and 3 **0**

2. 1. (covid-19 or coronavirus or 2019-ncov or sars-cov-2 or cov-19).mp. [mp=ti, ot, ab, tx, kw, ct, sh, hw]

2. ((((((family adj caregivers) or informal) adj caregivers) or unpaid) adj carers) or caregivers).mp. [mp=ti, ot, ab, tx, kw, ct, sh, hw]

3. unpaid adj car*).mp. [mp=ti, ot, ab, tx, kw, ct, sh, hw]

4. (((social adj isolation) or loneliness or quarantine or lockdown or social) adj dist*).mp. [mp=ti, ot, ab, tx, kw, ct, sh, hw]

5. 1 and 2 and 4 **0**

3. 1. (covid-19 or coronavirus or 2019-ncov or sars-cov-2 or cov-19).mp. [mp=ti, ot, ab, tx, kw, ct, sh, hw]

2. ((((((family adj caregivers) or informal) adj caregivers) or unpaid) adj carers) or caregivers).mp. [mp=ti, ot, ab, tx, kw, ct, sh, hw]

3. (((((((((caregiver adj burden) or caregiver) adj stress) or caregiver) adj fatigue) or car*) adj burnout) or car*) adj strain).mp. [mp=ti, ot, ab, tx, kw, ct, sh, hw]

4. 1 and 2 and 3 0

5. 1 and 3 0

**4****.** 1. (covid-19 or coronavirus or 2019-ncov or sars-cov-2 or cov-19).mp. [mp=ti, ot, ab, tx, kw, ct, sh, hw]

2. ((((((family adj caregivers) or informal) adj caregivers) or unpaid) adj carers) or caregivers).mp. [mp=ti, ot, ab, tx, kw, ct, sh, hw]

3. ((((((((mental adj health) and wellbeing) or well-being or well) adj being) or psychological) adj wellbeing) or well-being or well) adj being).mp. [mp=ti, ot, ab, tx, kw, ct, sh, hw]

4. 1 and 2 and 3 **4**

**Repeat search**

CINHAL Plus

**30/10/20 – limited to 2020/2021**

1. covid-19 or coronavirus or 2019-ncov or sars-cov-2 or cov-19 AND family caregivers or informal caregivers or unpaid carers or caregivers AND social isolation or loneliness or quarantine or lockdown **28**
2. covid-19 or coronavirus or 2019-ncov or sars-cov-2 or cov-19 AND family caregivers or informal caregivers or unpaid carers or caregivers AND mental health and wellbeing or well-being or well being OR psychological wellbeing or well-being or well being (Subject: Major Heading :covid-19**) 8**
3. covid-19 or coronavirus or 2019-ncov or sars-cov-2 or cov-19 AND family caregivers or informal caregivers or unpaid carers or caregivers AND caregiver burden or caregiver stress or caregiver fatigue or caregiver burnout or caregiver strain **14**
4. covid-19 or coronavirus or 2019-ncov or sars-cov-2 or cov-19 AND family caregivers or informal caregivers or unpaid carers or caregivers AND social distancing **11**
5. covid-19 or coronavirus or 2019-ncov or sars-cov-2 or cov-19 AND family caregivers or informal caregivers or unpaid carers or caregivers AND psychological stress or psychological distress **13**

**30/10/20 - ‘2020’ filter added**

APA PsycInfo 1806 to October Week 4 2020, Database Field Guide Embase 1974 to 2020 October 29, Database Field Guide Ovid MEDLINE(R) and Epub Ahead of Print, In-Process & Other Non-Indexed Citations and Daily - without Revisions 2016 to October 29, 2020

1. 1.(covid-19 or coronavirus or 2019-ncov or sars-cov-2 or cov-19).mp. [mp=ti, ab, hw, tc, id, ot, tm, mh, tn, dm, mf, dv, kw, fx, dq, nm, kf, ox, px, rx, an, ui, sy]

2. ((((((family adj caregivers) or informal) adj caregivers) or unpaid) adj carers) or caregivers).mp. [mp=ti, ab, hw, tc, id, ot, tm, mh, tn, dm, mf, dv, kw, fx, dq, nm, kf, ox, px, rx, an, ui, sy]

3. ((social adj isolation) or loneliness or quarantine or lockdown).mp. [mp=ti, ab, hw, tc, id, ot, tm, mh, tn, dm, mf, dv, kw, fx, dq, nm, kf, ox, px, rx, an, ui, sy]

4. (social adj distanc*).mp. [mp=ti, ab, hw, tc, id, ot, tm, mh, tn, dm, mf, dv, kw, fx, dq, nm, kf, ox, px, rx, an, ui, sy]

5. 3 or 4

6. 1 and 2 and 5 **198**

7. "2020".mp. [mp=ti, ab, hw, tc, id, ot, tm, mh, tn, dm, mf, dv, kw, fx, dq, nm, kf, ox, px, rx, an, ui, sy]

8. 6 and 7 **58**

**2.** 1. (covid-19 or coronavirus or 2019-ncov or sars-cov-2 or cov-19).mp. [mp=ti, ab, hw, tc, id, ot, tm, mh, tn, dm, mf, dv, kw, fx, dq, nm, kf, ox, px, rx, an, ui, sy]

2. ((((((family adj caregivers) or informal) adj caregivers) or unpaid) adj carers) or caregivers).mp. [mp=ti, ab, hw, tc, id, ot, tm, mh, tn, dm, mf, dv, kw, fx, dq, nm, kf, ox, px, rx, an, ui, sy]

3. ((((((mental adj health) and wellbeing) or well-being or well) adj being) or psychological wellbeing or well-being or well) adj being).mp. [mp=ti, ab, hw, tc, id, ot, tm, mh, tn, dm, mf, dv, kw, fx, dq, nm, kf, ox, px, rx, an, ui, sy]

4. 1 and 2 and 3 **40**

**3.** 1. (covid-19 or coronavirus or 2019-ncov or sars-cov-2 or cov-19).mp. [mp=ti, ot, ab, tx, kw, ct, sh, hw]

2. ((((((family adj caregivers) or informal) adj caregivers) or unpaid) adj carers) or caregivers).mp. [mp=ti, ot, ab, tx, kw, ct, sh, hw]

3. (((((((((caregiver adj burden) or caregiver) adj stress) or caregiver) adj fatigue) or car*) adj burnout) or car*) adj strain).mp. [mp=ti, ot, ab, tx, kw, ct, sh, hw]

4. 1 and 2 and 3 **2**

5. 1 and 3 **8**

**4.** 1. (covid-19 or coronavirus or 2019-ncov or sars-cov-2 or cov-19).mp. [mp=ti, ab, hw, tc, id, id, ot, tm, mh, tn, dm, mf, dv, kw, fx, dq, nm, kf, ox, px, rx, an, ui, sy]

2. ((((((family adj caregivers) or informal) adj caregivers) or unpaid) adj carers) or caregivers).mp. [mp=ti, ab, hw, tc, id, ot, tm, mh, tn, dm, mf, dv, kw, fx, dq, nm, kf, ox, px, rx, an, ui, sy]

3. ((mental adj health) or lonl*).mp. [mp=ti, ab, hw, tc, id, ot, tm, mh, tn, dm, mf, dv, kw, fx, dq, nm, kf, ox, px, rx, an, ui, sy]

4. 1 and 2 and 3 **92**

Cochrane Library via Evidence Based Medicine Reviews (EBMR) (Ovid Online)

1. 1. (covid-19 or coronavirus or 2019-ncov or sars-cov-2 or cov-19).mp. [mp=ti, ot, ab, tx, kw, ct, sh, hw]

2. ((((((family adj caregivers) or informal) adj caregivers) or unpaid) adj carers) or caregivers).mp. [mp=ti, ot, ab, tx, kw, ct, sh, hw]

3. ((((((mental adj health) and wellbeing) or well-being or well) adj being) or psychological) adj wel*).mp. [mp=ti, ot, ab, tx, kw, ct, sh, hw]

4. 1 and 2 and 3 **0**

2. 1. (covid-19 or coronavirus or 2019-ncov or sars-cov-2 or cov-19).mp. [mp=ti, ot, ab, tx, kw, ct, sh, hw]

2. ((((((family adj caregivers) or informal) adj caregivers) or unpaid) adj carers) or caregivers).mp. [mp=ti, ot, ab, tx, kw, ct, sh, hw]

3. unpaid adj car*).mp. [mp=ti, ot, ab, tx, kw, ct, sh, hw]

4. (((social adj isolation) or loneliness or quarantine or lockdown or social) adj dist*).mp. [mp=ti, ot, ab, tx, kw, ct, sh, hw]

5. 1 and 2 and 4 **1**

**02/11/20**

3. 1. (covid-19 or coronavirus or 2019-ncov or sars-cov-2 or cov-19).mp. [mp=ti, ot, ab, tx, kw, ct, sh, hw]

2. ((((((family adj caregivers) or informal) adj caregivers) or unpaid) adj carers) or caregivers).mp. [mp=ti, ot, ab, tx, kw, ct, sh, hw]

3. (((((((((caregiver adj burden) or caregiver) adj stress) or caregiver) adj fatigue) or car*) adj burnout) or car*) adj strain).mp. [mp=ti, ot, ab, tx, kw, ct, sh, hw]

4. 1 and 2 and 3 **0**

5. 1 and 3 **0**

**4.** 1. (covid-19 or coronavirus or 2019-ncov or sars-cov-2 or cov-19).mp. [mp=ti, ot, ab, tx, kw, ct, sh, hw]

2. ((((((family adj caregivers) or informal) adj caregivers) or unpaid) adj carers) or caregivers).mp. [mp=ti, ot, ab, tx, kw, ct, sh, hw]

3. ((((((((mental adj health) and wellbeing) or well-being or well) adj being) or psychological) adj wellbeing) or well-being or well) adj being).mp. [mp=ti, ot, ab, tx, kw, ct, sh, hw]

4. 1 and 2 and 3 **11**

**Appendix D - Grey literature search**

**Grey literature search: Websites**

**24/08/20**

**WHO**

Search 1 – Carers COVID 19

Search 2 **–** Caregivers and COVID 19

iSupport Lite - <https://www.who.int/teams/mental-health-and-substance-use/brain-health/integrated-care-support/isupport-lite>

Search 3**-** WHO in emergencies- Diseases – Outbreaks and crises- by disease- Coronavirus (COVID-19)

Overview of Public Health and Social Measures in the context of COVID-19 <https://www.who.int/publications/i/item/overview-of-public-health-and-social-measures-in-the-context-of-covid-19>

Search 4- Home/Emergencies/Diseases/Coronavirus disease (COVID-19)/Technical guidance publications- COVID-19: Risk communication and community engagement

-Mental health and psychosocial considerations during the COVID-19 outbreak

Interim guidance 18 March 2020 | COVID-19: Risk communication and community engagement

<https://www.who.int/publications/i/item/WHO-2019-nCoV-MentalHealth-2020.1>

**25/08/20**

**Rethink**

Covid 19 support – support for carers- 0

**Public Health England-** carers - 0

**CDC**- caregivers- 0

**MIND-** COVID 19 carers - 0

**CIRCLE Centre for International Research on Care, Labour & Equalities-**

CARING and COVID-19- Loneliness and use of services

<http://circle.group.shef.ac.uk/wp-content/uploads/2020/08/CARING-and-COVID-19-Loneliness-and-use-of-services_04.08.20.pdf>

Caring behind closed doors Forgotten families in the coronavirus outbreak: April 2020

<http://www.carersuk.org/images/News_and_campaigns/Behind_Closed_Doors_2020/Caring_behind_closed_doors_April20_pages_web_final.pdf>

**Euro carers**- links followed from CIRCLE

**(Researchers emailed 02/11/20**) <https://eurocarers.org/the-perspective-of-family-caregivers-on-the-impact-of-coronavirus-sars-cov-2-on-home-based-care-arrangements-in-germany-study-launched-in-germany/>

Covid-19 and care in Norway <https://eurocarers.org/covid-19-and-care-in-norway/>

**Family carers Ireland-** links followed from the Euro carers website search

CARING THROUGH COVID: LIFE IN LOCKDOWN <https://familycarers.ie/media/1394/caring-through-covid-life-in-lockdown.pdf>

**NHS England**

Carers, aging well, children and young people, Dementia, diabetes, integrated care, mental health and wheelchair services

HM Government, Department for Work and Pensions, Department for Digital, Culture, Media &Sport and Office for Civil Society and The Health foundation - no results.

**Carers UK** - for professionals – Policy and research

A Recovery Plan for carers <https://www.carersuk.org/for-professionals/policy/policy-library/a-recovery-plan-for-carers>

Policy and practice briefing: Improving carers’ access to food and carer ID <https://www.carersuk.org/for-professionals/policy/policy-library/policy-and-practice-briefing-improving-carers-access-to-food-and-carer-id>

Carers Week 2020 Research Report The rise in the number of unpaid carers during the coronavirus (COVID-19) outbreak <https://www.carersuk.org/images/CarersWeek2020/CW_2020_Research_Report_WEB.pdf>

Caring Behind Closed Doors: Forgotten Families in the Coronavirus Outbreak <https://www.carersuk.org/for-professionals/policy/policy-library/caring-behind-closed-doors-report>

**Carers Trust** - Resources

Coronavirus and the impact on caring <https://www.ons.gov.uk/peoplepopulationandcommunity/healthandsocialcare/conditionsanddiseases/articles/morepeoplehavebeenhelpingothersoutsidetheirhouseholdthroughthecoronaviruscovid19lockdown/2020-07-09>

No Longer Able to Care: Supporting older carers and ageing parent carers to plan for a future when they are less able or unable to care <https://carers.org/downloads/resources-pdfs/no-longer-able-to-care/no-longer-able-to-care.pdf>

**Google**

A search for full text of this research from the database searches: COVID-19 leaves unpaid carers without physical and mental health treatment led to ‘COVID-19 and Mental Health Current Awareness Bulletin 7^th^ August 2020’ where two relevant references were found:

Working with Older Caregivers of Persons with Mental Illness during COVID-19: Decreasing Burden, Creating Plans for Future Care, and Utilizing Strengths <https://pubmed.ncbi.nlm.nih.gov/32716263/>

One Month into the Reinforcement of Social Distancing due to the COVID-19 Outbreak: Subjective Health, Health Behaviors, and Loneliness among People with Chronic Medical Conditions <https://pubmed.ncbi.nlm.nih.gov/32727103/>

**Google scholar**

unpaid carers covid 19

Caregivers’ Mental Health and Somatic Symptoms During COVID-19 [**https://academic.oup.com/psychsocgerontology/advance-article/doi/10.1093/geronb/gbaa121/5879757**](https://academic.oup.com/psychsocgerontology/advance-article/doi/10.1093/geronb/gbaa121/5879757)

Canadian Geriatrics Society COVID-19 Recommendations for Older Adults. What Do Older Adults Need To Know?

**Repeated search**

**02/11/20**

**WHO**

Search 1 – Carers COVID 19 **0**

Search 2 – Caregivers and COVID 19 **0**

Search 3- WHO in emergencies- Diseases – Outbreaks and crises- by disease- Coronavirus (COVID-19) – Global research database search key word search: carers

- Development of a psychosocial intervention to support informal caregivers of people with end-stage kidney disease receiving haemodialysis <https://bmcnephrol.biomedcentral.com/articles/10.1186/s12882-020-02075-2>
- Impact of COVID-19 related social support service closures on people with dementia and unpaid carers: a qualitative study <https://www.tandfonline.com/doi/full/10.1080/13607863.2020.1822292>
- Exploring the ways in which COVID‐19 and lockdown has affected the lives of adult patients with anorexia nervosa and their carers <https://onlinelibrary.wiley.com/doi/10.1002/erv.2762>

Search 4**-** Home/Emergencies/Diseases/Coronavirus disease (COVID-19)/Technical guidance publications- COVID-19: Risk communication and community engagement- no results

**Rethink** – Covid 19 support – support for carers- 0

**Public Health England-** carers – 0

**05/11/20**

**CDC**- caregivers- 0

**MIND-** COVID 19 carers – 0

**CIRCLE Centre for International Research on Care, Labour & Equalities-** 0 new results

**Euro carers** – 0

**Family carers Ireland** – Publications- Research hub – 0 new results

**NHS England**

Carers, aging well, children and young people, Dementia, diabetes, integrated care, mental health and wheelchair services – searched from 24/08/20: no results

**Carers UK –** for professionals – Policy and research – no relevant reults

**Carers Trust** – Resources – no new results

**Google** - A repeat search for this research: COVID-19 leaves unpaid carers without physical and mental health treatment led to ‘COVID-19 and Mental Health Current Awareness Bulletin 7^th^ August 2020’ – no new results.

**Google scholar**

Impact of COVID-19 related social support service closures on people with dementia and unpaid carers: a qualitative study <https://www.tandfonline.com/doi/full/10.1080/13607863.2020.1822292>

Effect of the COVID‐19 pandemic on the mental health of carers of people with intellectual disabilities <https://onlinelibrary.wiley.com/doi/full/10.1111/jar.12811>

Impact of Covid-19 on the experiences of parents and family carers of autistic children and young people in the UK. UCL Research Briefing ID: 4992C01D-4415-480D-8088-341CF13EE1EB <https://discovery.ucl.ac.uk/id/eprint/10101297/3/Pavlopoulou_COVID19%20AUTISM%20FINAL%20GP.pdf>

**Google** – a search for: Impact of Covid-19 on the experiences of parents and family carers of autistic children and young people in the UK, led to a new resource: Left stranded: The impact of coronavirus on autistic people and their families in the UK <https://s4.chorus-mk.thirdlight.com/file/1573224908/63117952292/width=-1/height=-1/format=-1/fit=scale/t=444295/e=never/k=da5c189a/LeftStranded%20Report.pdf>

**Appendix E - Full list of excluded articles**

| Reason for exclusion | Study |
| --- | --- |
| Does not discuss impact on carers | Cardenas et al. (2020), Comas-Herrera et al. (2020), Eaton (2020), Ellerby (2020b), Eshraghi et al. (2020), Gray (2020), Kreimer (2020), Miller (2020), Mohile et al. (2020), Page et al. (2020), Senjam (2020), Saltzman et al. (2020) |
| Carers/caregivers refer to paid carers/healthcare workers | Ben-Pazi et al. (2020), Frankova (2020), Harrison and Webb (2020), Morley et al. (2020), Rao et al. (2020), Shah et al. (2020), Zhao et al. (2020) |
| Does not discuss or include carers | Montero-Odasso et al. (2020), Palmer (2020), Park et al. (2020), Passaro et al. (2020); Paterson (2020), Sibley et al. (2020), Tyrrell and Williams (2020); Who (2020a) |
| Article- not research | Acrbulletin (2020), Ellerby (2020a), Keng et al. (2020), Practicenurse (2020), Who (2020b), Who (2020d), Who (2020c) |
| Quantitative research | Consonni et al. (2020), Elran-Barak and Mozeikov (2020), Pagnini et al. (2020), Dhiman et al. (2020), Ergenekon et al. (2020); Gallagher and Wetherell (2020), Roy and Ayalon (2020), Russell et al. (2020); Zorcec et al. (2020) |
| Mixed method research | Chew et al. (2020), Hossain et al. (2020) |
| Letter/correspondence | Duong and Karlawish (2020), K. Chen (2020), Eckardt (2020), Rais et al. (2020), |
| Does not discuss mental health and wellbeing | Stokes and Patterson (2020) |
| Not about mental health and wellbeing of carers | Charles and Anderson-Nathe (2020), Swinford et al. (2020), Wang et al. (2020) |
| Hospital setting | Sutton-Smith (2020), Choi et al. (2020) |
| Not about carers | Aluh et al. (2020), Auerbach and Miller (2020), Banskota et al. (2020), Boldrini et al. (2020); L.-K. Chen (2020), Henning-Smith (2020), Irvine and Taylor (2020), Knopf (2020), Last (2020), Lei and Klopack (2020), Marini et al. (2020), Meskis (2020), Middleton (2020), Spinelli et al. (2020), Sun et al. (2020) |
| Caregivers refer to parents | Prime et al. (2020), Russell et al. (2020) |
| Focus on people who have been COVID 19 positive | Brown et al. (2020) |
| Full text unavailable | Worldofirishnursing (2020) |
| Conference abstract | Cona et al. (2020), Harbegue et al. (2020) |
| comment | Radbruch et al. (2020), Steinman et al. (2020) |

http://ovidsp.ovid.com/ovidweb.cgi?T=JS&PAGE=reference&D=emexb&NEWS=N&AN=632276254

https://www.ncbi.nlm.nih.gov/pmc/articles/PMC7332840/pdf/fneur-11-00746.pdf (Accessed: 18th August 2020).

Boldrini, P., Kiekens, C., Bargellesi, S., Brianti, R., Galeri, S., Lucca, L., Montis, A., Posteraro, F., Scarponi, F., Straudi, S. & Negrini, S. (2020). First impact of COVID-19 on services and their preparation. "Instant paper from the field" on rehabilitation answers to the COVID-19 emergency. *European journal of physical and rehabilitation medicine* [Online], 56(3)**,** pp. 319-322. Available at: https://www.minervamedica.it/en/journals/europa-medicophysica/article.php?cod=R33Y2020N03A0319 (Accessed: 18th August 2020).

Brown, C., Peck, S., Humphreys, J., Schoenherr, L., Saks, N. T., Sumser, B. & Elia, G. (2020). COVID-19 Lessons: The Alignment of Palliative Medicine and Trauma-Informed Care. *Journal of Pain and Symptom Management* [Online], 60(2)**,** pp. e26-e30. Available at: http://www.elsevier.com/locate/jpainsymman (Accessed: 18th August 2020).

Cardenas, M. C., Bustos, S. S. & Chakraborty, R. (2020). A 'Parallel Pandemic': The Psychosocial Burden of Covid-19 in Children and Adolescents. *Acta paediatrica (Oslo, Norway : 1992)* [Online]. Available at: http://ovidsp.ovid.com/ovidweb.cgi?T=JS&PAGE=reference&D=medp&NEWS=N&AN=32799388 (Accessed: 18th August 2020).

Charles, G. & Anderson-Nathe, B. (2020). Uncertainty in the Time of Coronavirus. *Child & Youth Services* [Online], 41(1)**,** pp. 1-2. Available at: https://blogs.scientificamerican.com/observations/uncertainty-in-a-time-of-coronavirus/ (Accessed: 2nd August 2020).

Chen, K. (2020). 'Use of Gerontechnology to Assist Older Adults to Cope with the COVID-19 Pandemic', *Journal of the American Medical Directors Association,* 21(7), pp. 983-984.

Chen, L.-K. (2020). Older adults and COVID-19 pandemic: Resilience matters. *Archives of Gerontology & Geriatrics* [Online], 89**,** pp. N.PAG-N.PAG. Available at: https://www.ncbi.nlm.nih.gov/pmc/articles/PMC7247489/ (Accessed: 10th August 2020).

Chew, Q. H., Wei, K. C., Vasoo, S., Chua, H. C. & Sim, K. (2020). 'Narrative synthesis of psychological and coping responses towards emerging infectious disease outbreaks in the general population: practical considerations for the COVID-19 pandemic', *Singapore medical journal,* 61(7), pp. 350-356.

Choi, K. R., Heilemann, M. V., Fauer, A. & Mead, M. (2020). A Second Pandemic: Mental Health Spillover From the Novel Coronavirus (COVID-19). 26**,** pp. 340-343. Available at: https://journals.sagepub.com/doi/full/10.1177/1078390320919803 (Accessed: 11th August 2020).

Comas-Herrera, A., Fernandez, J.-L., Hancock, R., Hatton, C., Knapp, M., McDaid, D., Malley, J., Wistow, G. & Wittenberg, R. (2020). COVID-19: Implications for the Support of People with Social Care Needs in England. *Journal of Aging & Social Policy* [Online], 32(4/5)**,** pp. 365-372. Available at: https://www.tandfonline.com/doi/full/10.1080/08959420.2020.1759759 (Accessed: 20th August 2020).

Cona, M. S., Dalu, D., Ferrario, S., Tosca, N., Gambaro, A. R., Filipazzi, V., Rota, S. & La Verde, N. M. (2020). The emotional impact of COVID-19 outbreak on cancer out-patients and their caregivers: Impressions from the heart of the Italian pandemic. *Annals of Oncology* [Online], 31(Supplement 4)**,** pp. S956-S957. Available at: https://www.annalsofoncology.org/article/S0923-7534(20)42049-6/fulltext (Accessed: 9th November 2020).

Consonni, M., Telesca, A., Dalla Bella, E., Bersano, E. & Lauria, G. (2020). Amyotrophic lateral sclerosis patients' and caregivers' distress and loneliness during COVID-19 lockdown. *Journal of neurology* [Online]. Available at: https://www.ncbi.nlm.nih.gov/pmc/articles/PMC7372539/ (Accessed: 5th September 2020).

Dhiman, S., Sahu, P. K., Reed, W. R., Ganesh, G. S., Goyal, R. K. & Jain, S. (2020). Impact of COVID-19 outbreak on mental health and perceived strain among caregivers tending children with special needs. *Research in developmental disabilities* [Online], 107**,** p. 103790. Available at: https://www.sciencedirect.com/science/article/pii/S0891422220302225?via%3Dihub (Accessed: 9th November 2020).

Duong, M. T. & Karlawish, J. (2020). Caregiving at a physical distance: Initial thoughts for COVID-19 and beyond. *Journal of the American Geriatrics Society* [Online], 68(6)**,** pp. 1170-1172. Available at: https://onlinelibrary.wiley.com/doi/10.1111/jgs.16495 (Accessed: 9th November 2020).

Eaton, J. (2020). Wellbeing and mental health during the COVID-19 outbreak. *Community Eye Health Journal* [Online], 33(109)**,** pp. 5-6. Available at: https://cehjournal.org/wp-content/uploads/CEHJ109_Covid-19_MentalHealth.pdf (Accessed: 20th November 2020).

Eckardt, J. P. (2020). Caregivers of people with severe mental illness in the COVID-19 pandemic. *The Lancet Psychiatry* [Online], 7(8)**,** p. e53. Available at: https://www.thelancet.com/journals/lanpsy/article/PIIS2215-0366(20)30252-2/fulltext (Accessed: 27th September 2020).

Ellerby, K. (2020a). Caring for carers: Everyday we see patients who would struggle to get through their daily life without the support of someone else to help them. In the absence of professional or voluntary organisational support, who is there within the community to enable these people to live their lives? Do we look at our patients in the wider context and ask the question -- who is actually caring for them? *Practice Nurse* [Online], 50(4)**,** pp. 24-28. Available at: http://www.practicenurse.co.uk/index.php?p1=articles&p2=2050 (Accessed: 9th November 2020).

Ellerby, K. (2020b). Caring for vulnerable patients during the COVID-19 pandemic: With more than two million patients categorised as extremely vulnerable, what should general practice nurses be doing to ensure they are provided with necessary care? *Practice Nurse* [Online], 50(5)**,** pp. 19-23. Available at: http://www.practicenurse.co.uk/index.php?p1=articles&p2=2064 (Accessed: 10th August 2020).

Elran-Barak, R. & Mozeikov, M. (2020). One Month into the Reinforcement of Social Distancing due to the COVID-19 Outbreak: Subjective Health, Health Behaviors, and Loneliness among People with Chronic Medical Conditions. *International journal of environmental research and public health* [Online], 17(15)**,** p. 5403. Available at:

https://www.ncbi.nlm.nih.gov/pmc/articles/PMC7432045/ (Accessed: 20th November 2020).

Ergenekon, A. P., Yegit, C. Y., Cenk, M., Ikizoglu, N. B., Atag, E., Gokdemir, Y., Eralp, E. E. & Karadag, B. (2020). Depression and Anxiety in Mothers of Home Ventilated Children Before and During COVID-19 Pandemic. *Pediatric pulmonology* [Online]. Available at: https://onlinelibrary.wiley.com/doi/10.1002/ppul.25107 (Accessed: 9th November 2020).

Eshraghi, A. A., Li, C., Alessandri, M., Messinger, D. S., Eshraghi, R. S., Mittal, R. & Armstrong, F. D. (2020). 'COVID-19: Overcoming the challenges faced by individuals with autism and their families', *The Lancet Psychiatry,* 7(6), pp. 481-483.

Frankova, H. (2020). The impact of COVID-19 on people with autism, learning disabilities and mental health conditions. *Nursing & Residential Care* [Online], 22(6)**,** pp. 1-3. Available at: https://www.magonlinelibrary.com/doi/full/10.12968/nrec.2020.22.6.10 (Accessed: 30th August 2020).

Gallagher, S. & Wetherell, M. A. (2020). Risk of depression in family caregivers: unintended consequence of COVID-19. *BJPsych open* [Online], 6(6)**,** p. e119. Available at: https://www.cambridge.org/core/journals/bjpsych-open/article/risk-of-depression-in-family-caregivers-unintended-consequence-of-covid19/65EAC3BD84DAE8EDE57701E169BE92A9 (Accessed: 10th November 2020).

Gray, V. (2020). Protecting the elderly through and beyond the Covid-19 lockdown. *Journal of Community Nursing* [Online], 34(3)**,** pp. 12-13. Available at: http://search.ebscohost.com/login.aspx?direct=true&AuthType=ip,shib&db=jlh&AN=143663229&site=ehost-live (Accessed:

Harbegue, K., Mejri, N., El Benna, H., Berrazega, Y., Rachdi, H., Labidi, S. & Boussen, H. (2020). COVID-19: An additional burden on caregivers of cancer patients in Tunisia. *Annals of Oncology* [Online], 31(Supplement 4)**,** p. S1017. Available at: https://www.annalsofoncology.org/article/S0923-7534(20)41798-3/fulltext (Accessed: 10th November 2020).

Harrison, D. & Webb, J. O. (2020). MOVING AND HANDLING SOLUTIONS IN RESPONSE TO THE COVID-19 PANDEMIC IN THE UK: EXPLORING THE IMPACT OF COVID-19 ON A CAREGIVER'S ABILITY TO CARRY OUT THEIR ROLE IN HOME CARE. *International Journal of Safe Patient Handling & Mobility (SPHM)* [Online], 10(2)**,** pp. 55-58. Available at: https://sphmjournal.com/product/moving-handling-solutions-response-covid-19-pandemic-uk-exploring-impact-covid-19-caregivers-ability-carry-role-home-care/ (Accessed: 30th August 2020).

Henning-Smith, C. (2020). The Unique Impact of COVID-19 on Older Adults in Rural Areas. *Journal of Aging & Social Policy* [Online], 32(4/5)**,** pp. 396-402. Available at: https://www.tandfonline.com/doi/full/10.1080/08959420.2020.1770036 (Accessed: 22nd August 2020).

Hossain, M. M., Sultana, A. & Purohit, N. (2020). Mental health outcomes of quarantine and isolation for infection prevention: a systematic umbrella review of the global evidence. *Epidemiology & Health* [Online], 42**,** pp. e2020038-e2020038. Available at: https://www.ncbi.nlm.nih.gov/pmc/articles/PMC7644933/ (Accessed: 1st September 2020).

Irvine, H. & Taylor, J. (2020). 'Prioritising workload during the pandemic: It has been said that general practice has been transformed more dramatically in the past few weeks than in the previous decade, but the rapid pace of change may leave some nurses in doubt about what they should -- and should not -- be doing', *Practice Nurse,* 50(5), pp. 7-12.

Keng, A., Brown, E. E., Rostas, A., Rajji, T. K., Pollock, B. G., Mulsant, B. H. & Kumar, S. (2020). Effectively Caring for Individuals With Behavioral and Psychological Symptoms of Dementia During the COVID-19 Pandemic. *Frontiers in Psychiatry* [Online], 11**,** p. 573367. Available at: http://www.frontiersin.org/Psychiatry (Accessed: 10th November 2020).

Knopf, A. (2020). 'During and after COVID‐19, anxiety and depression will increase: Study', *Brown University Child & Adolescent Behavior Letter,* 36(9), pp. 6-7.

Kreimer, S. (2020). Neuropalliative Care During COVID-19--Addressing Isolation, Fear, and Terminal Illness. *Neurology Today* [Online], 20(11)**,** pp. 21-22. Available at: https://journals.lww.com/neurotodayonline/Fulltext/2020/06040/Neuropalliative_Care_During_COVID_19_Addressing.10.aspx (Accessed: 20th August 2020).

Last, R. (2020). 'Lifestyle, health and wellbeing - COVID-19 and beyond', *Practice Nurse,* 50(6), pp. 22-25.

Lei, M.-K. & Klopack, E. T. (2020). 'Social and Psychological Consequences of the COVID-19 Outbreak: The Experiences of Taiwan and Hong Kong', *Psychological Trauma: Theory, Research, Practice & Policy,* 12, pp. S35-S37.

Marini, C. M., Pless Kaiser, A., Smith, B. N. & Fiori, K. L. (2020). 'Aging Veterans' Mental Health and Well-Being in the Context of COVID-19: The Importance of Social Ties During Physical Distancing', *Psychological Trauma: Theory, Research, Practice & Policy,* 12, pp. S217-S219.

Meskis, S. (2020). 'COPING THROUGH THE PANDEMIC', *Alaska Nurse,* 71(2), pp. 14-16.

Middleton, K. (2020). 'Look after yourselves', *Frontline (20454910),* 26(5), pp. 38-38.

Miller, E. A. (2020). 'Protecting and Improving the Lives of Older Adults in the COVID-19 Era', *Journal of Aging & Social Policy,* 32(4/5), pp. 297-309.

Mohile, S., Dumontier, C., Mian, H., Loh, K. P., Williams, G. R., Wildes, T. M., Boyd, K., Ramsdale, E., Pyne, S., Magnuson, A., Tew, W., Klepin, H. D., Dale, W. & Shahrokni, A. (2020). 'Perspectives from the Cancer and Aging Research Group: Caring for the vulnerable older patient with cancer and their caregivers during the COVID-19 crisis in the United States', *Journal of Geriatric Oncology,* 11(5), pp. 753-760.

Montero-Odasso, M., Goens, S. D., Kamkar, N., Lam, R., Madden, K., Molnar, F., Speechley, M. & Stranges, S. (2020). 'Canadian Geriatrics Society COVID-19 Recommendations for Older Adults. What Do Older Adults Need To Know?', *Canadian Geriatrics Journal,* 23(1), pp. 149-151.

Morley, G., Sese, D., Rajendram, P. & Horsburgh, C. C. (2020). 'Addressing caregiver moral distress during the COVID-19 pandemic', *Cleveland Clinic journal of medicine*.

Page, N., Naik, V., Singh, P., Fernandes, P., Nirabhawane, V. & Chaudhari, S. (2020). 'Homecare and the COVID-19 pandemic – Experience at an urban specialist cancer palliative center', *Indian Journal of Palliative Care,* 26, pp. 63-69.

Pagnini, F., Bonanomi, A., Tagliabue, S., Balconi, M., Bertolotti, M., Confalonieri, E., Di Dio, C., Gilli, G., Graffigna, G., Regalia, C., Saita, E. & Villani, D. (2020). 'Knowledge, Concerns, and Behaviors of Individuals During the First Week of the Coronavirus Disease 2019 Pandemic in Italy', *JAMA Network Open,* 3(7), pp. e2015821-e2015821.

Palmer, N. (2020). Vulnerability of patient, families and carers in a climate of COVID-19. *Journal of Kidney Care* [Online], 5(3)**,** pp. 138-139. Available at: https://www.magonlinelibrary.com/doi/full/10.12968/jokc.2020.5.3.138 (Accessed: 1st September 2020).

Park, C. L., Russell, B. S., Fendrich, M., Finkelstein-Fox, L., Hutchison, M. & Becker, J. (2020). Americans' COVID-19 Stress, Coping, and Adherence to CDC Guidelines. *Journal of General Internal Medicine* [Online], 35(8)**,** pp. 2296-2303. Available at: https://link.springer.com/article/10.1007%2Fs11606-020-05898-9 (Accessed: 26th September 2020).

Passaro, A., Addeo, A., Von Garnier, C., Blackhall, F., Planchard, D., Felip, E., Dziadziuszko, R., de Marinis, F., Reck, M., Bouchaab, H. & Peters, S. (2020). 'ESMO Management and treatment adapted recommendations in the COVID-19 era: Lung cancer', *ESMO open,* 5(Supplement 3).

Paterson, R. (2020). 'Long-term effects of COVID-19: impact on prescribing practice', *Journal of Prescribing Practice,* 2(6), pp. 274-275.

PracticeNurse (2020). Survey measures negative impact of COVID-19 on mental health. *Practice Nurse* [Online], 50(6)**,** pp. 9-9. Available at: http://search.ebscohost.com/login.aspx?direct=true&AuthType=ip,shib&db=jlh&AN=144273011&site=ehost-live (Accessed: 11th November 2020).

Prime, H., Wade, M. & Browne, D. T. (2020). 'Risk and Resilience in Family Well-Being During the COVID-19 Pandemic', *American Psychologist,* 75(5), pp. 631-643.

Radbruch, L., Knaul, F. M., de Lima, L., de Joncheere, C. & Bhadelia, A. (2020). The key role of palliative care in response to the COVID-19 tsunami of suffering. *The Lancet* [Online], 395(10235)**,** pp. 1467-1469. Available at: https://www.thelancet.com/journals/lancet/article/PIIS0140-6736(20)30964-8/fulltext (Accessed: 10th November 2020).

Rais, N. C., Au, L. & Tan, M. (2020). 'COVID-19 Impact in Community Care—A Perspective on Older Persons With Dementia in Singapore', *Journal of the American Medical Directors Association,* 21(7), pp. 997-997.

Rao, S., Spruijt, O., Sunder, P., Daniel, S., Chittazhathu, R., Nair, S., Leng, M., M M, S., Raghavan, B., Manuel, A., Rijju, V., Vijay, G., Prabhu, A., Parameswaran, U. & Venkateswaran, C. (2020). 'Psychosocial aspects of COVID-19 in the context of palliative care – A quick review', *Indian Journal of Palliative Care,* 26, pp. 116-120.

Roy, S. & Ayalon, L. (2020). "Goodness and kindness": Long distance caregiving through volunteers during the COVID-19 lockdown in India. *The journals of gerontology. Series B, Psychological sciences and social sciences* [Online]. Available at: https://academic.oup.com/psychsocgerontology/advance-article/doi/10.1093/geronb/gbaa187/5942527 (Accessed: 10th November 2020).

Russell, B. S., Hutchison, M., Tambling, R., Tomkunas, A. J. & Horton, A. L. (2020). Initial Challenges of Caregiving During COVID-19: Caregiver Burden, Mental Health, and the Parent-Child Relationship. *Child psychiatry and human development* [Online], 51(5)**,** pp. 671-682. Available at: https://link.springer.com/article/10.1007%2Fs10578-020-01037-x (Accessed: 10th November 2020).

Saltzman, L. Y., Hansel, T. C. & Bordnick, P. S. (2020). 'Loneliness, Isolation, and Social Support Factors in Post-COVID-19 Mental Health', *Psychological Trauma: Theory, Research, Practice & Policy,* 12, pp. S55-S57.

Senjam, S. S. (2020). 'Impact of COVID-19 pandemic on people living with visual disability', *Indian journal of ophthalmology,* 68(7), pp. 1367-1370.

Shah, K., Bedi, S., Onyeaka, H., Singh, R. & Chaudhari, G. (2020). 'The Role of Psychological First Aid to Support Public Mental Health in the COVID-19 Pandemic', *Cureus,* 12(6), p. e8821.

Sibley, C. G., Greaves, L. M., Satherley, N., Wilson, M. S., Overall, N. C., Lee, C. H. J., Milojev, P., Bulbulia, J., Osborne, D., Milfont, T. L., Houkamau, C. A., Duck, I. M., Vickers-Jones, R. & Barlow, F. K. (2020). 'Effects of the COVID-19 Pandemic and Nationwide Lockdown on Trust, Attitudes Toward Government, and Well-Being', *American Psychologist,* 75(5), pp. 618-630.

Spinelli, M., Lionetti, F., Pastore, M. & Fasolo, M. (2020). 'Parents' Stress and Children's Psychological Problems in Families Facing the COVID-19 Outbreak in Italy', *Frontiers in psychology,* 11, p. 1713.

Steinman, M. A., Perry, L. & Perissinotto, C. M. (2020). Meeting the Care Needs of Older Adults Isolated at Home During the COVID-19 Pandemic. *JAMA internal medicine* [Online], 180(6)**,** pp. 819-820. Available at: https://jamanetwork.com/journals/jamainternalmedicine/fullarticle/2764748 (Accessed: 10th November 2020).

Stokes, J. E. & Patterson, S. E. (2020). Intergenerational Relationships, Family Caregiving Policy, and COVID-19 in the United States. *Journal of Aging & Social Policy* [Online], 32(4/5)**,** pp. 416-424. Available at: https://www.tandfonline.com/doi/full/10.1080/08959420.2020.1770031 (Accessed: 26th August 2020).

Sun, S., Lin, D. & Operario, D. (2020). 'Need for a Population Health Approach to Understand and Address Psychosocial Consequences of COVID-19', *Psychological Trauma: Theory, Research, Practice & Policy,* 12, pp. S25-S27.

Sutton-Smith, L. (2020). 'Planning for a COVID-19 crisis', *Kai Tiaki Nursing New Zealand,* 26(4), pp. 26-27.

Swinford, E., Galucia, N. & Morrow-Howell, N. (2020). Applying gerontological social work perspectives to the coronavirus pandemic. *Journal of Gerontological Social Work* [Online]**,** pp. No-Specified. Available at: https://www.tandfonline.com/doi/full/10.1080/01634372.2020.1766628 (Accessed: 21st August 2020).

Tyrrell, C. J. & Williams, K. N. (2020). 'The Paradox of Social Distancing: Implications for Older Adults in the Context of COVID-19', *Psychological Trauma: Theory, Research, Practice & Policy,* 12, pp. S214-S216.

Wang, T., Liu, S., Joseph, T. & Lyou, Y. (2020). 'Managing bladder cancer care during the COVID-19 pandemic using a team-based approach', *Journal of Clinical Medicine,* 9(5), p. 1574.

WHO (2020a). Guidance on COVID-19 for the care of older people and people living in long-term care facilities, other non-acute care facilities and home care. (WPR/DSE/2020/015). Available at: http://iris.wpro.who.int/handle/10665.1/14500 (Accessed: 20th August 2020).

WHO (2020b). iSupport Lite. (23rd September 2020). Available at: https://www.who.int/teams/mental-health-and-substance-use/brain-health/integrated-care-support/isupport-lite (Accessed: 26th September 2020).

WHO (2020c). Mental health and psychosocial considerations during the

COVID-19 outbreak Available at: https://www.who.int/publications/i/item/WHO-2019-nCoV-MentalHealth-2020.1 (Accessed: 23rd September 2020).

WHO (2020d). Overview of Public Health and Social Measures in the context of COVID-19. Available at: https://www.who.int/publications/i/item/overview-of-public-health-and-social-measures-in-the-context-of-covid-19 (Accessed: 26th September 2020).

WorldofIrishNursing (2020). Maintaining services for people with rare diseases. *World of Irish Nursing & Midwifery* [Online]**,** pp. 49-49. Available at: https://inmo.ie/tempDocs/FULL%20ISSUE%20FEB%202017.pdf (Accessed: 18th August 2020).

Zhao, F., Ahmed, F. & Faraz, N. A. (2020). Caring for the caregiver during COVID-19 outbreak: Does inclusive leadership improve psychological safety and curb psychological distress? A cross-sectional study. *International journal of nursing studies* [Online], 110**,** p. 103725. Available at: https://www.sciencedirect.com/science/article/pii/S002074892030211X?via%3Dihub (Accessed: 10th November 2020).

Zorcec, T., Jakovska, T., Micevska, V., Boskovska, K. & Cholakovska, V. C. (2020). Pandemic with COVID-19 and Families with Children with Chronic Respiratory Diseases. *Prilozi (Makedonska akademija na naukite i umetnostite. Oddelenie za medicinski nauki)* [Online], 41(2)**,** pp. 95-101. Available at: https://www.sciencedirect.com/science/article/pii/S002074892030211X?via%3Dihub (Accessed: 10th November 2020).

**Appendix 7 – List of excluded records after eligibility assessment**

| Reason for exclusion | Study |
| --- | --- |
| Quantitative research | Carersuk (2020b), Carersuk (2020a), Cohen et al. (2020); Eurocarers (2020), Giebel et al. (2020), Ons (2020), Park (2020), Prasad et al. (2020), Willner et al. (2020) |
| Mixed method | Alves et al. (2020), Centre for International Research on Care (2020), Familycarersireland (2020), Ng et al. (2020) |
| Policy | Carersuk (2020d), Carersuk (2020c) |
| Case report | Gulia et al. (2020) |
| Article | Greenberg et al. (2020); Payne (2020) |
| Commentary | Kent et al. (2020), Migliaccio and Bouzigues (2020) |
| Letter/correspondence | Labrum et al. (2020), Sannes et al. (2020) |
| Not about carers | Lai et al. (2020), Padala et al. (2020) |
| Not about COVID 19 | Carerstrust (2020) |
| Failed appraisal, | Dhavale et al. (2020), |
| Does not discuss mental health and wellbeing of carers | Canevelli et al. (2020) |

Alves, G. S., Casali, M. E., Veras, A. B., Carrilho, C. G., Bruno Costa, E., Rodrigues, V. M. & Dourado, M. C. N. (2020). A Systematic Review of Home-Setting Psychoeducation Interventions for Behavioral Changes in Dementia: Some Lessons for the COVID-19 Pandemic and Post-Pandemic Assistance. *Frontiers in Psychiatry* [Online], 11**,** p. 577871. Available at: https://www.frontiersin.org/articles/10.3389/fpsyt.2020.579985/full (Accessed: 10th November 2020).

Canevelli, M., Valletta, M., Toccaceli Blasi, M., Remoli, G., Sarti, G., Nuti, F., Sciancalepore, F., Ruberti, E., Cesari, M. & Bruno, G. (2020). Facing dementia during the COVID-19 outbreak. *Journal of the American Geriatrics Society* [Online], 68(8)**,** pp. 1673-1676. Available at: https://onlinelibrary.wiley.com/doi/full/10.1111/jgs.16644 (Accessed: 10th November 2020).

CarersTrust (2020). 'No Longer Able to Care: Supporting older carers and ageing parent carers to plan for a future when they are less able or unable to care'.

CarersUK (2020a). Carers Week 2020 Research Report: The rise in the number of unpaid carers during the coronavirus (COVID-19) outbreak. Available at: https://www.carersuk.org/images/CarersWeek2020/CW_2020_Research_Report_WEB.pdf (Accessed: 25th August 2020).

CarersUK (2020b). Caring behind closed doors Forgotten families in the coronavirus outbreak: April 2020. Available at: http://www.carersuk.org/images/News_and_campaigns/Behind_Closed_Doors_2020/Caring_behind_closed_doors_April20_pages_web_final.pdf (Accessed: 24th September 2020).

CarersUK (2020c). Policy and practice briefing: Improving carers’ access to food and carer ID Available at: https://www.carersuk.org/for-professionals/policy/policy-library/policy-and-practice-briefing-improving-carers-access-to-food-and-carer-id (Accessed: 24th September 2020).

CarersUK (2020d). A Recovery Plan for carers Available at: https://www.carersuk.org/for-professionals/policy/policy-library/a-recovery-plan-for-carers (Accessed: 24th September 2020).

Centre for International Research on Care, L. E. (2020). CARING and COVID-19- Loneliness and use of services Available at: http://circle.group.shef.ac.uk/wp-content/uploads/2020/08/CARING-and-COVID-19-Loneliness-and-use-of-services_04.08.20.pdf (Accessed: 30th August 2020).

Cohen, G., Russo, M. J., Campos, J. A. & Allegri, R. F. (2020). Living with dementia: increased level of caregiver stress in times of COVID-19. *International psychogeriatrics* [Online]**,** pp. 1-5. Available at: https://www.cambridge.org/core/journals/international-psychogeriatrics/article/living-with-dementia-increased-level-of-caregiver-stress-in-times-of-covid19/529C561EDCE2211D2C8345F15ED50763 (Accessed: 10th November 2020).

Dhavale, P., Koparkar, A. & Fernandes, P. (2020). Palliative care interventions from a social work perspective and the challenges faced by patients and caregivers during COVID-19. *Indian Journal of Palliative Care* [Online], 26**,** pp. 58-62. Available at: http://search.ebscohost.com/login.aspx?direct=true&AuthType=ip,shib&db=jlh&AN=144403282&site=ehost-live (Accessed: 1st November 2020).

EuroCarers (2020). Covid-19 and care in Norway. Available at: https://eurocarers.org/covid-19-and-care-in-norway/ (Accessed: 24th Septmember 2020).

FamilyCarersIreland (2020). CARING THROUGH COVID: LIFE IN LOCKDOWN. Available at: https://familycarers.ie/media/1394/caring-through-covid-life-in-lockdown.pdf (Accessed: 24th September 2020).

Giebel, C., Lord, K., Cooper, C., Shenton, J., Cannon, J., Pulford, D., Shaw, L., Gaughan, A., Tetlow, H., Butchard, S., Limbert, S., Callaghan, S., Whittington, R., Rogers, C., Komuravelli, A., Rajagopal, M., Eley, R., Watkins, C., Downs, M., Reilly, S., Ward, K., Corcoran, R., Bennett, K. & Gabbay, M. (2020). A UK survey of COVID-19 related social support closures and their effects on older people, people with dementia, and carers. *International journal of geriatric psychiatry* [Online]. Available at: https://onlinelibrary.wiley.com/doi/10.1002/gps.5434 (Accessed: 10th November 2020).

Greenberg, N. E., Wallick, A. & Brown, L. M. (2020). Impact of COVID-19 pandemic restrictions on community-dwelling caregivers and persons with dementia. *Psychological trauma : theory, research, practice and policy* [Online], 12(S1)**,** pp. S220-S221. Available at: https://doi.apa.org/fulltext/2020-43961-001.html (Accessed: 20th September 2020).

Gulia, A., Mishra, S. & Bhatnagar, S. (2020). Multiple caregiving role with the novel challenge of COVID-19 pandemic: A crisis situation. *Indian Journal of Palliative Care* [Online], 26**,** pp. 163-165. Available at: http://www.jpalliativecare.com/article.asp?issn=0973-1075;year=2020;volume=26;issue=5;spage=163;epage=165;aulast=Gulia (Accessed: 20th September 2020).

Kent, E. E., Ornstein, K. A. & Dionne-Odom, J. N. (2020). The Family Caregiving Crisis Meets an Actual Pandemic. *Journal of Pain & Symptom Management* [Online], 60(1)**,** pp. e66-e69. Available at: https://www.jpsmjournal.com/article/S0885-3924(20)30203-7/fulltext (Accessed: 24th September 2020).

Labrum, T., Newhill, C. & Smathers, T. (2020). Working with Older Caregivers of Persons with Mental Illness during COVID-19: Decreasing Burden, Creating Plans for Future Care, and Utilizing Strengths. *Journal of Gerontological Social Work* [Online]**,** pp. 1-5. Available at: https://pubmed.ncbi.nlm.nih.gov/32716263/ (Accessed: 25th September 2020).

Lai, F. H. Y., Yan, E. W. H., Yu, K. K. Y., Tsui, W. S., Chan, D. T. H. & Yee, B. K. (2020). The Protective Impact of Telemedicine on Persons With Dementia and Their Caregivers During the COVID-19 Pandemic. *American Journal of Geriatric Psychiatry* [Online]. Available at: https://www.sciencedirect.com/science/article/pii/S1064748120304383?via%3Dihub (Accessed: 10th November 2020).

Migliaccio, R. & Bouzigues, A. (2020). Dementia and COVID-19 Lockdown: More Than a Double Blow for Patients and Caregivers. *Journal of Alzheimer's Disease Reports* [Online], 4(1)**,** pp. 231-235. Available at: https://content.iospress.com/articles/journal-of-alzheimers-disease-reports/adr200193 (Accessed: 28th September 2020).

Ng, K. Y. Y., Zhou, S., Tan, S. H., Ishak, N. D. B., Goh, Z. Z. S., Chua, Z. Y., Chia, J. M. X., Chew, E. L., Shwe, T., Mok, J. K. Y., Leong, S. S., Lo, J. S. Y., Ang, Z. L. T., Leow, J. L., Lam, C. W. J., Kwek, J. W., Dent, R., Tuan, J., Lim, S. T., Hwang, W. Y. K., Griva, K. & Ngeow, J. (2020). Understanding the Psychological Impact of COVID-19 Pandemic on Patients With Cancer, Their Caregivers, and Health Care Workers in Singapore. *JCO global oncology* [Online], 6**,** pp. 1494-1509. Available at: https://ascopubs.org/doi/10.1200/GO.20.00374 (Accessed: 10th November 2020).

ONS (2020). Coronavirus and the impact on caring. Available at: https://www.ons.gov.uk/peoplepopulationandcommunity/healthandsocialcare/conditionsanddiseases/articles/morepeoplehavebeenhelpingothersoutsidetheirhouseholdthroughthecoronaviruscovid19lockdown/2020-07-09 (Accessed: 28th September 2020).

Padala, K. P., Wilson, K. B., Gauss, C. H., Stovall, J. D. & Padala, P. R. (2020). VA Video Connect for Clinical Care in Older Adults in a Rural State During the COVID-19 Pandemic: Cross-Sectional Study. *Journal of Medical Internet Research* [Online], 22(9)**,** pp. N.PAG-N.PAG. Available at: https://www.ncbi.nlm.nih.gov/pmc/articles/PMC7537724/ (Accessed: 10th November 2020).

Park, S. S. (2020). Caregivers' Mental Health and Somatic Symptoms During Covid-19. *The journals of gerontology. Series B, Psychological sciences and social sciences* [Online]. Available at: http://ovidsp.ovid.com/ovidweb.cgi?T=JS&PAGE=reference&D=emexb&NEWS=N&AN=632503362 (Accessed: 26th September 2020).

Payne, J. (2020). EMOTIONAL HEALTH IN ISOLATION: SUPPORTING SOMEONE WITH DISABILITIES AT HOME DURING COVID-19. *Exceptional Parent* [Online], 50(7)**,** pp. 44-45. Available at: https://reader.mediawiremobile.com/epmagazine/issues/206228/viewer?page=45 (Accessed: 28th September 2020).

Prasad, S., Holla, V., Neeraja, K., Surisetti, B., Kamble, N., Yadav, R., Pal, P., Holla, V. V., Surisetti, B. K. & Pal, P. K. (2020). Impact of Prolonged Lockdown due to COVID-19 in Patients with Parkinson's Disease. *Neurology India* [Online], 68(4)**,** pp. 792-795. Available at: https://pubmed.ncbi.nlm.nih.gov/32859814/ (Accessed: 10th November 2020).

Sannes, T. S., Yeh, I. M. & Gray, T. F. (2020). Caring for loved ones with cancer during the COVID-19 pandemic: A double hit risk for social isolation and need for action. *Psycho-Oncology* [Online], 29(9)**,** pp. 1418-1420. Available at: http://onlinelibrary.wiley.com/journal/10.1002/(ISSN)1099-1611 (Accessed: 28th September 2020).

Willner, P., Rose, J., Stenfert Kroese, B., Murphy, G. H., Langdon, P. E., Clifford, C., Hutchings, H., Watkins, A., Hiles, S. & Cooper, V. (2020). Effect of the COVID-19 pandemic on the mental health of carers of people with intellectual disabilities. *Journal of Applied Research in Intellectual Disabilities* [Online], 33(6)**,** pp. 1523-1533. Available at: https://onlinelibrary.wiley.com/doi/10.1111/jar.12811 (Accessed: 1st November 2020).

**Appendix G – Full critical appraisals**

| Study 1 | Dhavale et al. (2020) |
| --- | --- |
| 1.Was there a clear statement of the aims of the research? | **Yes** - The authors provide a rationale for the research which is followed by 2 clear aims. |
| 2.Is qualitative  methodology  appropriate? | **Yes** - The research aims to understand the experiences of palliative care patients and their caregivers during the COVID 19 pandemic. It also explores the impact of the interventions used by social workers to support the patients and their caregivers. |
| 3.Was the research design appropriate to address the aims of the research? | **Can’t Tell** - The authors do not discuss their rationale for using case notes as their methodology for their research or what their research protocol was. |
| 4.Was the recruitment strategy appropriate to the aims of the research? | **Can’t Tell** - The authors have given a brief description of the participants included in the research; however, they do not provide clear inclusion/exclusion criteria or discuss their selection process. Nine families were selected to participate in the research with no further information, for example what stage of their cancer trajectory they are at or where they are receiving their care. |
| 5.Was the data collected in  a way that addressed the  research issue? | **Can’t Tell** - Data collection is not discussed beyond the source of information |
| 6. Has the relationship  between researcher and  participants been  adequately considered? | **No** - The role of the authors is not made clear beyond an affiliation with Cipla Palliative Care and Training Center. |
| 7. Have ethical issues been taken into consideration? | **Can’t Tell** - Ethical considerations are not discussed in this article, such as how consent was obtained, how the research was explained or if ethical approval was sought. The authors have declared no conflicts of interest. |
| 8. Was the data analysis sufficiently rigorous? | **No** - A framework approach was used to develop themes however this this is the extent of the information provided on data analysis |
| 9. Is there a clear statement of findings? | **Can’t Tell** - The findings are presented in a clear manner and clearly address the research question divided into three categories. However, the authors do not discuss the credibility of their findings and the lack of information in the data collection and analysis section makes it difficult to make an assessment of the findings |
| 10. How valuable is the  research? | **Weak** - Transferability to other settings is not discussed by the authors, neither are specific recommendations/suggestions for further research. There is a lack of information on the research design, recruitment strategy and data collection which reduce the transferability of the research |

| Study 2 | Vaitheswaran et al. (2020) |
| --- | --- |
| 1.Was there a clear statement of the aims of the research? | **Yes** - The authors provide a clear objective of the research which is expanded and clearly explained in the introduction section. |
| 2.Is qualitative  methodology  appropriate? | **Yes** - The research aims to describe the experiences and needs of caregivers of people who have dementia, during the COVID 19 pandemic. Therefore, a qualitative research method is appropriate to address their research goal. |
| 3.Was the research design appropriate to address the aims of the research? | **Yes** - The authors have not explained their decision to use telephone interviews, however they have explained that the research was carried out during a period of lockdown; it is understandable that face to face interviews would not be possible. Semi-structured interview questions were used to provide an opportunity for people to describe their experiences in detail. |
| 4.Was the recruitment strategy appropriate to the aims of the research? | **Yes** - The recruitment strategy is clearly explained and purposive sampling was used. The limitation paragraph points out that only people who were contactable by phone were included and the research was carried out in one center. Recruitment was stopped when data saturation was reached at 31 participants out of 56 which is appropriate. |
| 5.Was the data collected in  a way that addressed the  research issue? | **Can’t Tell** -. No information is provided about the questions which were asked or if a topic guide was used during the semi-structured interview. The interviews were transcribed although the authors do not explicitly say how the interviews were recorded. There is no mention of any modifications to the research methods. Three researchers coded the transcripts independently, this could have been expanded on to include information about the outcome of the coding process, for example were there any disagreements? |
| 6. Has the relationship  between researcher and  participants been  adequately considered? | **Can’t tell** - The authors state that telephone interviews were conducted by consultants who had previously interacted with the patients and their care givers during appointments. There is no discussion of how this may or may not influence the research. There is also no clarification of the roles, each researcher holds besides the role they played in the research. The limitation section does consider that the use of one service to carry out the research could affect the results. |
| 7. Have ethical issues been taken into consideration? | **Can’t Tell** - Ethical approval for the research was granted by the Institutional Ethics Committee at the Schizophrenia Research Foundation. There is no discussion about how the research was explained or how consent was obtained, especially as caregivers would be sharing information about the people they care for as well as themselves. All participants have been anonymised, however, there is no discussion of how this was explained to the participants or whether any participants expressed any concerns about participating. The participants were given the opportunity to express their suggestions of what could be done to address their support needs. The authors have declared no conflict of interest. |
| 8. Was the data analysis sufficiently rigorous? | **Can’t tell** - The authors provide a brief description of the data analysis process, explaining that a thematic analysis approach was used to collate codes into identifiable categories, it would have been useful to provide more information such as categories which were formed before a consensus was reached between the researchers. There is also no mention of how many researchers were involved in the data analysis process after the three researchers transcribed and coded the data. Once the categories were decided several quotations are used to evidence the relevance of the themes developed, however there is a lack of information on the process leading to the final themes and there is no discussion of any views which may have opposed the final themes. There is little evidence of reflexivity from the researchers as they have not discussed their roles, relationships with the participants or how their views could potentially be a source of bias. |
| 9. Is there a clear statement of findings? | **Yes** - The findings are presented using quotations and discussions of the themes identified during data analysis, although there is little discussion of any alternative views. More than one person analysed the data; however it was not specified how many people were involved in analysing the data. As there is a lack of information of the categories before a consensus was reached, respondent validation would have increased the credibility of their findings. The findings are discussed in the context of the original research question and culminate in the authors suggesting that lessons should be learnt from the pandemic so that systems have more capacity to support patients and caregivers when faced with additional challenges. |
| 10. How valuable is the  research? | **Medium** - This research adds valuable data to an area of research which as yet has a paucity of information and evidence. The researchers identify existing policies in India and add suggestions to improve support for unpaid carers and the people they care for. Although the authors do not discuss transferability, their research design and recruitment strategy provide information which can be used to assess transferability in other settings. There is information about the participants and the systems in place in their country which can also be used to assess transferability to a UK setting. |

| Study 3 | Savla et al. (2020) |
| --- | --- |
| 1.Was there a clear statement of the aims of the research? | **Yes –** There is sparse information about the impact of the COVID 19 pandemic on caregivers and their coping strategies |
| 2.Is qualitative  methodology  appropriate? | **Yes** – the authors aim to assess the subjective accounts from caregivers |
| 3.Was the research design appropriate to address the aims of the research? | **Can’t tell** – The authors do not discuss the reasons they chose their research design. However, as the research was carried out during a period of lockdown it is clear that telephone interviews are an appropriate research design as they are able to obtain relevant information with no physical contact. |
| 4.Was the recruitment strategy appropriate to the aims of the research? | **Can’t tell** – the study was carried out in two stages: at the start of the stay-at-home order and two weeks after it started. Although the authors mention where recruitment was carried out, there is no information about how or why the participants were recruited. There is more information given in stage two of the research |
| 5.Was the data collected in  a way that addressed the  research issue? | **Can’t tell** – 30-minute interviews using survey questions and open ended questions: there is no information about the questions that were asked ie. was everyone asked the same questions, what questions were asked, was more or less time needed for any of the interviews, what were the survey items? |
| 6. Has the relationship  between researcher and  participants been  adequately considered? | **No** – the authors explain their role in the research; however they do not discuss any relationship between them and the participants or how their own views or experiencing could potentially influence their interpretation of the study results. The declare no conflict of interest. |
| 7. Have ethical issues been taken into consideration? | **Can’t tell** – Consent was obtained from the participants, however, there is not mention of how this was done, if ethical approval for the research was sought or granted or how the research was explained to the participants. |
| 8. Was the data analysis sufficiently rigorous? | **Can’t tell** – the authors provide a summary of the data analysis process; more information is needed. |
| 9. Is there a clear statement of findings? | **Yes** – relevant to the research aim |
| 10. How valuable is the  research? | **Weak**– the authors highlight that a small research sample was used in a specific area: rural Virginia and so the results may not be generalisable. However, the research provides some insight into the coping mechanisms of caregivers during the first two weeks of the stay-at-home order. Unfortunately, the authors do not conclude with specific recommendations although they do suggest home and internet-based delivery of services as well as improved internet access in rural areas. |

| Study 4 | Giebel et al. (2020) |
| --- | --- |
| 1.Was there a clear statement of the aims of the research? | **Yes**- exploration of the impact of reduced access to social care and support services on people with dementia and their carers during the COVID 19 outbreak. Background information is given to explain the relevance of the research. |
| 2.Is qualitative  methodology  appropriate? | **Yes**- the research aims to explore the experience of people who have dementia and their carers from their perspective. |
| 3.Was the research design appropriate to address the aims of the research? | **Can’t tell** – The authors do not discuss the reasons they chose their research design. However, as the research was carried out during a period of lockdown it is clear that telephone interviews are an appropriate research design as they can obtain relevant information with no physical contact. |
| 4.Was the recruitment strategy appropriate to the aims of the research? | **Yes**- the authors provide a detailed explanation of their recruitment strategy which used convenience sampling. They also acknowledge that due to the need to gather information quickly they imposed a time limit to recruitment which may have had an impact on the number and type of participants involved i.e., more carers were interested in participating than people who had dementia who had mental capacity to express their interest and consent. |
| 5.Was the data collected in  a way that addressed the  research issue? | **Yes**- a clear explanation of the data collection process is given including how the topic guide was developed, how long the semi-structured interviews lasted, examples of the type of questions asked, who conducted the interviews as well as their experience levels and how the interviews were recorded and transcribed. |
| 6. Has the relationship  between researcher and  participants been  adequately considered? | **yes**- the authors involved people living with dementia, carers, academic team member and practitioners in the development of the interview topic guide. 2 researchers used inductive thematic analysis to code up to 35 transcripts discussing their results until agreed themes were generated which were used to code the remaining transcripts. The research team included 1 person living with dementia and 3 former unpaid carers. |
| 7. Have ethical issues been taken into consideration? | **Yes** – the research was explained to participants initially at the start of recruitment and then over the phone. Verbal consent was obtained and ethical approval was granted through the University of Liverpool. The involvement of a person living with dementia and 3 former unpaid carers aimed to ensure the study, interpretation and implication of findings were grounded in the lived experiences of those affected by dementia. |
| 8. Was the data analysis sufficiently rigorous? | **Yes**- a detailed description of the data analysis process is given. audio recordings were transcribed verbatim by experienced typists and read several times by the research team. inductive thematic analysis was used for up to 35 transcripts and deductive thematic analysis for the remaining transcripts which resulted in 3 major themes. |
| 9. Is there a clear statement of findings? | **Yes**- the authors discuss their findings in detail which are relevant to the research aim. They conclude with recommendations to policy makers. |
| 10. How valuable is the  research? | **Strong** – this research provides valuable insight to the experience of both carers and people living with dementia and the access to services during the COVID 19 pandemic. Although it is not the focus of the research, the mental health and wellbeing of carers is also discussed as a consequence of the measures in place to address the current pandemic. |

| Study 5 | Clark Bryan et al. (2020) |
| --- | --- |
| 1.Was there a clear statement of the aims of the research? | **Yes-** exploration of the impact of the COVID 19 outbreak on the lives of adult patients with anorexia nervosa and their carers |
| 2.Is qualitative  methodology  appropriate? | **Yes-** the research aims to explore the experiences of patients and their carers during the COVID 19 pandemic |
| 3.Was the research design appropriate to address the aims of the research? | **Yes**- the authors explain how they developed the research using recruits from an existing Randomised Controlled Trial (RCT) to gain insight from the participants during the pandemic. This was not the purpose of the original and ongoing RCT research; however, it will contribute to the ongoing trial. |
| 4.Was the recruitment strategy appropriate to the aims of the research? | **Yes**- the authors explain the recruitment process and include information about participants who were excluded giving reasons. The participants are described giving age ranges and roles of their carers i.e., parent/partner |
| 5.Was the data collected in  a way that addressed the  research issue? | **Yes**- the authors state the question used in a semi-structured interview with the participants over skype which was recorded and transcribed verbatim. Follow up questions were asked in cases where more information was needed. Saturation of data was reached at 49 participants. However, the setting of data collection is not made clear. |
| 6. Has the relationship  between researcher and  participants been  adequately considered? | **No**- the authors do not discuss their roles or possible sources of bias which may occur in their research. They adapted their research design to be both relevant to their research aim and to the ongoing RCT in the form of qualitative information. |
| 7. Have ethical issues been taken into consideration? | **Can’t tell** – as the research participants are recruited from an existing trial which their research will contribute to, there is implied ethical approval. However, this is not discussed by the authors. The participants have given consent for this research although there is no discussion of how consent was obtained or what was explained to the participants. The authors have given clear explanations of how data was anonomysed to maintain confidentiality |
| 8. Was the data analysis sufficiently rigorous? | **Yes-** thematic analysis was used and carried out by two researchers independently with joint discussions on five occasions until themes were agreed on. The data analysis process is clearly explained and the results are reported clearly |
| 9. Is there a clear statement of findings? | **Yes**- the findings are relevant to the research aim and are explained using quotations including quotations which are in the minority of other participants views/experiences. Themes are presented and discussed in detail. The strengths and limitations of the research are also considered and explained |
| 10. How valuable is the  research? | **Strong** – Although the research is aimed at patients of anorexia nervosa and their carers, the themes which have emerged contribute to an evolving evidence base of the impact of the COVID 19 measure put in place to address the pandemic, on unpaid carers and the people they care for. The themes are also clearly separated for patients and carers. |

**Appendix H – Characteristics of included studies**

| Author | Aim of study | Domain of care needs | Geographical context | Participants | Methodology | Data collection | Results |
| --- | --- | --- | --- | --- | --- | --- | --- |
| Vaitheswaran et al. (2020) | - description of the experiences and needs of people who care for family members who have dementia in India during the COVID 19 pandemic. | dementia | India-  outpatient clinic | 31 of 56 family caregivers.  Data saturation reached at 31 participants | qualitative | semi-structured telephone interviews | thematic analysis |
| Bbc and Sounds (2020) | - experience of becoming a carer during the COVID 19 pandemic | Alzheimer’s disease – 1  multiple sclerosis  and dementia – 1  Alzheimer’s and dementia – 1  Alzheimer’s and vascular dementia -1  Physical care needs- 1  severe learning disability, autism, and severe heart defect – 1  stroke – 1  dementia- 1  intravenous feeds - 1 | England- individuals’ homes | 7 of 8, 1 excluded due to person requiring care being a care home resident  -1 caring for a parent and spouse  -2 caring for 1 parent  -1 caring for both parents  -1 caring for a spouse  1- caring for a partner  -1 caring for an adult child | qualitative -on a radio phone in | telephone interview |  |
| Clark Bryan et al. (2020) | - impact of COVID 19 and associated lockdown measures on adult patients with anorexia nervosa and their carers | anorexia nervosa | UK- 15 specialist units across the UK | 21 patients who have anorexia nervosa.  28 carers of the patients. Of the 28 carers 3 were partners and 25 were parents. Data saturation reached at 49 participants. | qualitative | semi-structured interviews using skype |  |
| Giebel et al. (2020) | - exploration of the impact of reduced access to social care and support services on people with dementia and their carers during the COVID 19 outbreak | dementia | England – North West coast and nationally | 42 unpaid carers, 8 people living with dementia.  55% of carers were spouses.  5 carers were caring for someone who lived in a care home. | qualitative | semi-structured interviews over the phone,  topic guides used |  |

**Appendix I- Descriptive themes**

| Concerns | New routines | Financial worries | Access to support | What will happen after the pandemic |
| --- | --- | --- | --- | --- |
| Inability to recognise symptoms of COVID 19 in relatives who cannot verbally express their needs, Hospitalisation of relative | Change in daily routines/activities | Unable to go to work | Specialists | Social care support |
| Quality of life | Diet and levels of physical activity for relative |  | Medications | Access to services |
| Exacerbation of existing or development of new health and behaviour problems | Increased levels of care needs |  | Respite | Changes in policies |
| Health of carer | Diet and levels of physical activity for carer |  | Ideas of activities at home |  |
| Breaking lockdown rules | Reduced support |  | Care packages |  |
| protecting relatives from getting COVID-19 | Working from home |  |  |  |
|  | Care delivery |  |  |  |
|  | Increased carer responsibility |  |  |  |
|  | Reduced pressure/more time at home |  |  |  |
|  | Reflection |  |  |  |

**Appendix J - Analytical themes**

| Immediate worries | Adapting to change | Post pandemic fears | Use of technology |
| --- | --- | --- | --- |
| Health and/or behaviour deterioration | Routines and/or activities | Social care support | Access to services |
| Protection from exposure to COVID 19 | Reduced physical activity of people who require care |  | Care delivery |
| Health of carer | Reduced support |  |  |
|  | Increased carer responsibility |  |  |

**Appendix K – Frequency charts**

**Immediate worries/fears**

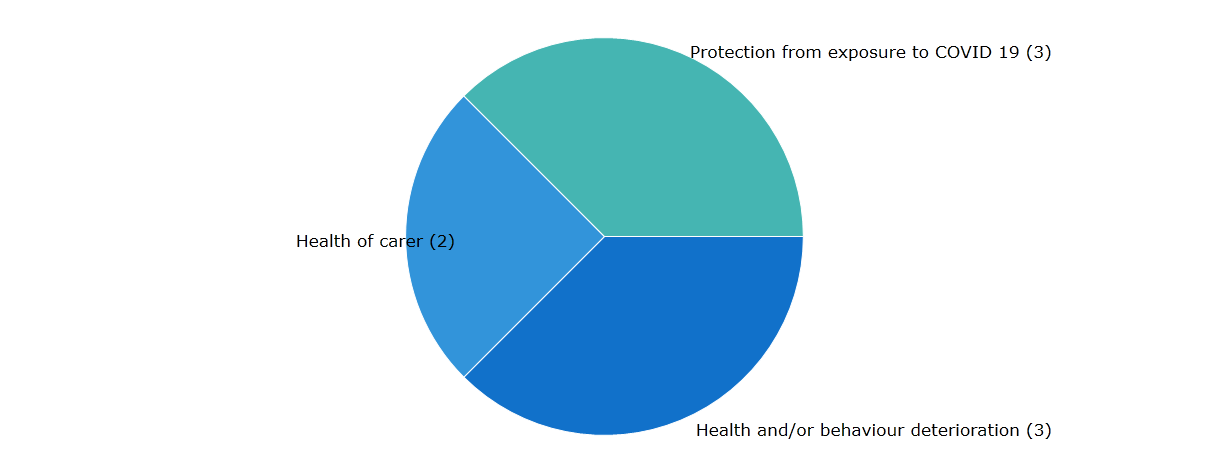

**Adapting to change**

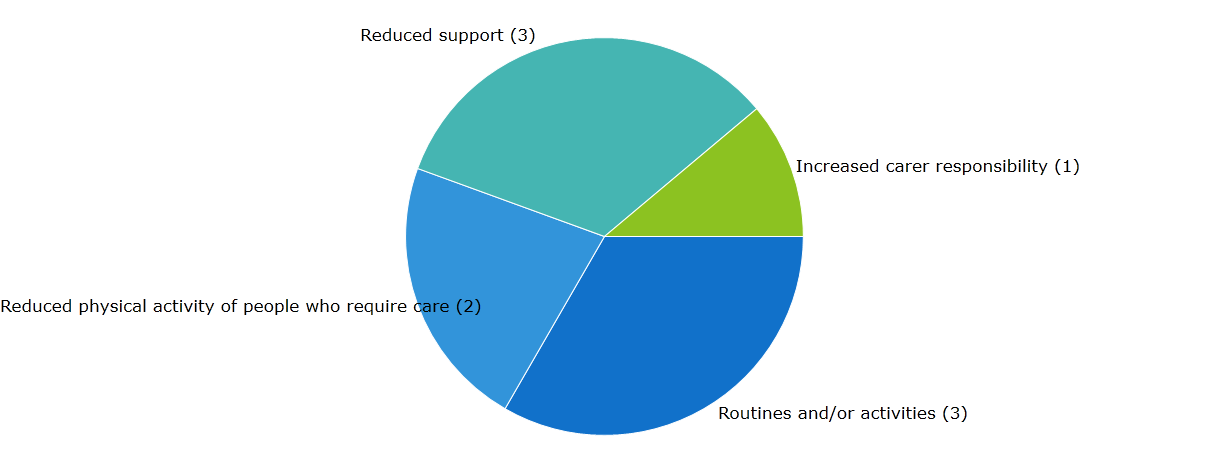

**Post pandemic fears**

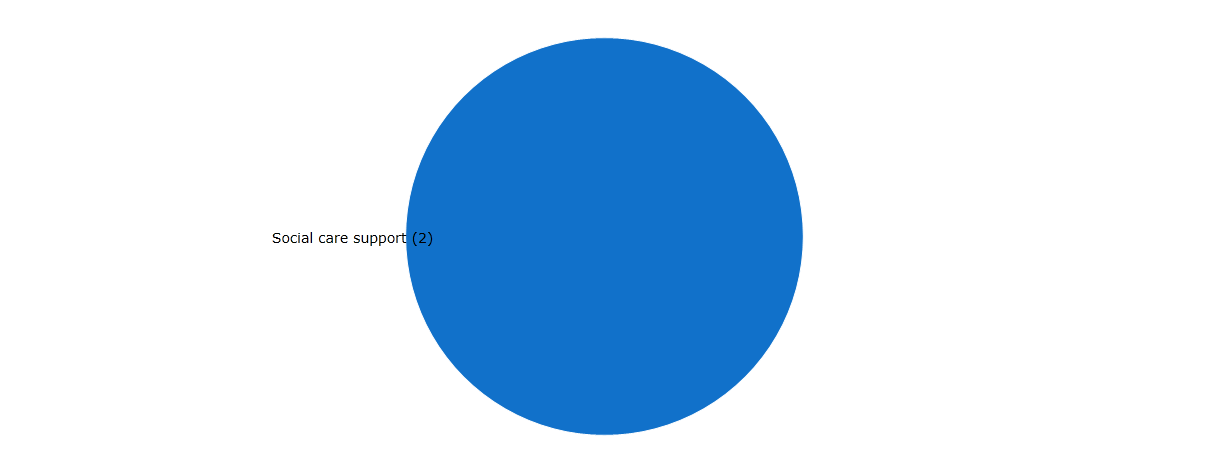

**Use of Technology**

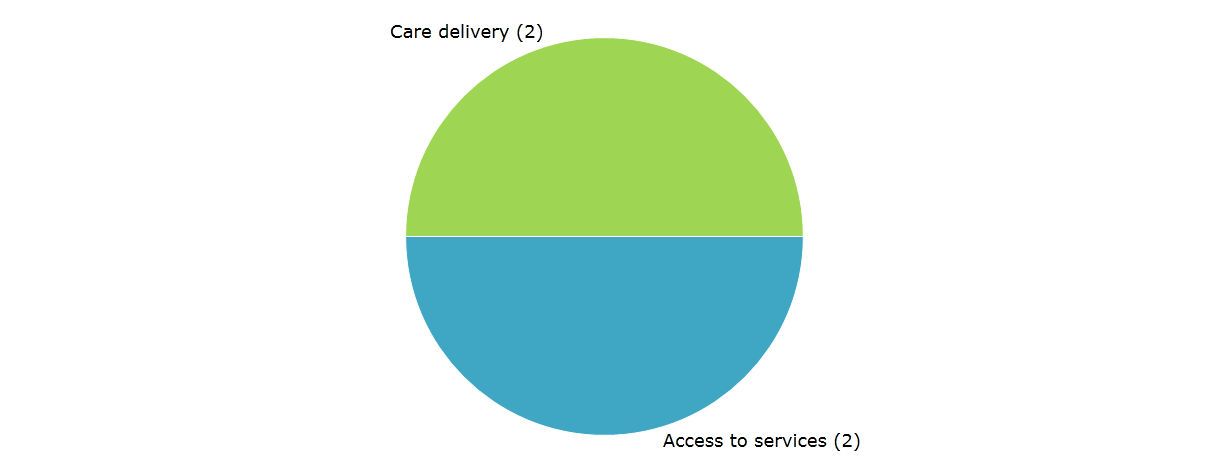
